# Pregnancy and neonatal outcomes of COVID-19 – co-reporting of common outcomes from the PAN-COVID and AAP SONPM registry

**DOI:** 10.1101/2021.01.06.21249325

**Authors:** Edward Mullins, Mark L. Hudak, Jayanta Banerjee, Trace Getzlaff, Julia Townson, Kimberly Barnette, Rebecca Playle, Tom Bourne, Christoph Lees, On behalf of PAN-COVID-investigators, National Perinatal COVID-19 Registry Study Group

## Abstract

**Background:** Few large, cohort studies report data on individual’s maternal, fetal, perinatal, and neonatal outcomes associated with SARS-CoV-2 infection in pregnancy. We report outcomes from a collaboration formed early during the pandemic between the investigators of two registries, the UK and global Pregnancy and Neonatal outcomes in COVID-19 (PAN-COVID) study and the US American Academy of Pediatrics Section on Neonatal Perinatal Medicine (AAP SONPM) National Perinatal COVID-19 Registry.

**Methods:** PAN-COVID (suspected or confirmed SARS-CoV-2 infection at any stage in pregnancy) and the AAP SONPM registry (positive maternal testing for SARS-CoV-2 from 14 days before delivery to 3 days after delivery) studies collected data on maternal, fetal, perinatal and neonatal outcomes. PAN-COVID results are presented as all inclusions and those with confirmed SARS-CoV-2 infection only.

**Results:** We report 4004 women in pregnancy affected by suspected or confirmed SARS-CoV-2 infection (1606 from PAN-COVID and 2398 from the AAP SONPM) from January 1^st^ 2020 to July 25^th^ 2020 (PAN-COVID) and August 8^th^ (AAP SONPM). For obstetric outcomes in PAN-COVID and AAP SONPM, respectively, maternal death occurred in 0.5% and 0.17%, early neonatal death in 0.2% and 0.3%, and stillbirth in 0.50% and 0.65% of women. Delivery was pre-term (<37 weeks gestation) in 12% of all women in PAN-COVID, in 16.2% of those women with confirmed infection in PAN-COVID and 16.2% of women in AAP SONPM. Very preterm delivery (< 27 weeks’ gestation) occurred in 0.6% in PAN-COVID and 0.7% in AAP SONPM.

Neonatal SARS-CoV-2 infection was reported in 0.8% of PAN-COVID all inclusions, 2.0% in PAN-COVID confirmed infections and 1.8% in the AAP SONPM study; the proportions of babies tested were 9.5%, 20.7% and 87.2% respectively.

The proportion of SGA babies was 8.2% in PAN-COVID all inclusions, 9.7% in PAN-COVID confirmed infection and 9.6% in AAP SONPM. Gestational age adjusted mean z-scores were −0.03 for PAN-COVID and −0.18 for AAP SONPM.

**Conclusions:** The findings from the UK and US SARS-CoV-2 in pregnancy registries were remarkably concordant. Pre-term delivery affected a higher proportion of women in pregnancy than expected from historical and contemporaneous national data. The proportions of women affected by stillbirth, small for gestational age infants and early neonatal death were comparable to historical and contemporaneous UK and US data. Although maternal death was uncommon, the proportion was higher than expected from UK and US population data, likely explained by under-ascertainment of women affected by milder and asymptomatic infection in pregnancy. The data presented support strong guidance for enhanced precautions to prevent SARS-COV-2 infection in pregnancy, particularly in the context of increased risks of preterm delivery and maternal mortality, and for priority vaccination of women planning pregnancy.

**What is known about SARS-COV-2 infection in pregnancy and neonates?:** Cohort, population surveillance studies and living systematic reviews have included limited numbers of women in pregnancy affected by COVID-19 and report that most women and infants had good outcomes.

**What this study adds:** Preterm deliveries occurred in a high proportion of women participating in these two registries in comparison to contemporaneous and historical national data in the UK and US. The majority of preterm deliveries occurred late preterm (between 32+0 and 36+6 weeks’ gestation).

SARS-COV-2 infection in pregnancy did not appear to be associated with a clinically significant effect on the rate of stillbirth, fetal growth, or neonatal outcomes.

Although maternal death was uncommon, the proportion was higher than expected from UK and US population data, likely explained by under-ascertainment of women affected by milder and asymptomatic infection in pregnancy.

## Background

At the onset of the COVID-19 pandemic, WHO designated pregnant women as a vulnerable group based on preliminary reports of increased risk of stillbirth, preterm birth and fetal growth restriction (FGR) and extrapolation from experience with previous pandemic respiratory viruses including SARS, MERS ^1,2,3^ and influenza. SARS-CoV-2 outcome data derived predominantly from case series have variably reported diagnostic testing, maternal, fetal and neonatal outcomes, and transmission to the neonate^4^. Clinical outcomes appeared worse for pregnant compared with non-pregnant women infected with SARS^5,6^ and H1N1 influenza^1,7^. Pregnant women were assumed to be at heightened risk of morbidity and mortality from SARS-CoV-2 infection, particularly when symptomatic^8^.

Current knowledge about the effect of SARS-CoV-2 infection in pregnancy has been largely gathered from case reports, case series and population surveillance systems in high income countries. These data have focused particularly on maternal outcomes of women with symptomatic disease and report data on maternal death, stillbirths and neonatal deaths of around 1% ^9^ in this context. The risk of an infant testing positive for SARS-CoV-2 is approximately 2.5% in women admitted to hospital with symptomatic disease^9^. A strong case for vertical transmission has been reported in one neonate ^10^. A living systematic review and meta-analysis suggests that pregnant women with symptomatic SARS-CoV-2 infection are less likely to present with fever and myalgia and more likely to receive intensive care, to require ventilation, and to experience a higher risk of preterm birth^1,2^. In low and middle income countries (LMIC), the impact may have been greater. For instance, the rate of stillbirth was increased by a third and neonatal mortality threefold in one Nepalese cohort^11^.

Centre-based registries gather case data prospectively on the effect of SARS-CoV-2 infection from healthcare systems around the world and offer both a practical and scalable method to accrue clinical outcomes on key research questions from a variety of populations and healthcare systems. Two pregnancy registries in English speaking countries have analogous aims and similar design. The UK’s Pregnancy and Neonatal Outcomes of COVID-19 (PAN-COVID) study utilizes a dataset that collected outcome data focussed on determining the effect of SARS-CoV-2 infection on miscarriage, fetal growth restriction, stillbirth, pre-term delivery, vertical transmission (suspected or confirmed) and early-onset symptomatic neonatal SARS-CoV-2 infection and includes fields on ultrasound diagnosis and neonatal care not included in other more general studies (Banerjee BMJ Open 2020). The study has 177 centres participating in the UK and 10 countries around the world. The American Academy of Pediatrics (AAP) Section on Neonatal Perinatal Medicine (SONPM) National Perinatal COVID-19 Registry collects data on women who have tested positive for SARS-CoV-2 on samples obtained from 14 days before to 3 days after delivery. The study has 288 centres across the United States. Both registries collect maternal demographic and symptomatology data as well as perinatal and neonatal outcomes.

## Methods

**The PAN-COVID study** used a purpose-built Elsevier Macro database and collected outcome data from pregnant women with confirmed or suspected SARS-CoV-2 infection who delivered between 1/1/2020 and 31/3/2021 and their neonates. The study was sponsored by Imperial College London and funded by UK Research Institute (UKRI) and NIHR. The PAN-COVID study is detailed in the published protocol (Banerjee BMJ Open), and we briefly describe the methodology below. The main study objectives for PAN-COVID were to establish a UK and international disease registry for women with confirmed or suspected SARS-CoV-2 infection in pregnancy. Women with suspected SARS-CoV-2 infection were included because capacity for SARS-CoV-2 PCR testing in the UK was limited to hospital-admitted patients only until April 2020 and we expected the majority of women with infection not to receive testing. Suspected SARS-CoV-2 infection was in a untested pregnant woman when she reported symptoms that her healthcare professionals thought were likely due to SARS-CoV-2.

We focused on the following research questions: in women recruited to the PAN-COVID registry with confirmed or suspected SARS-CoV-2 infection, what were the incidences of miscarriage, fetal growth restriction, stillbirth, pre-term birth and transmission to the fetus or neonate by vertical or other routes of infection. Recruitment was by verbal consent and retrospective inclusion was permitted. Weekly contact open sessions were co-ordinated by NW London CRN and the study management team to motivate staff to recruit and enter data. The UK CRN support network of research midwives was facilitated by Urgent Public Health research designation. The study was registered with ISRCTN (ISRCTN68026880).

**The AAP SONPM National Perinatal COVID-10 Registry** collect data via a REDCap database from pregnant women who tested positive for SARS-COV-2 on specimens obtained between 14 days before delivery to 3 days after delivery and opened on 4/1/2020. The study was funded by the University of Florida and in-kind contributions from participating centers. This registry rapidly accrued detailed pregnancy, perinatal and neonatal outcome data from mother/infant dyads to assess the impact of SARS-CoV-2 infection across a range of US healthcare settings.

### Outcomes

#### PAN-COVID

Assessment of outcomes required follow-up by individual healthcare professionals who accessed medical records routinely available to them as part of the clinical care team. When a pregnant woman with suspected or confirmed SARS-CoV-2 infection was registered on the database, they were automatically assigned with a unique participant identification number. The limit for data collection was 28 days after the delivery or pregnancy loss of the last woman registered.

#### AAP SONPM National Registry

Participating investigators extracted outcome data from the hospital electronic medical record and electronically transmitted de-identified data to a REDCap data base housed on a secure University of Florida server.

### Statistical analysis

#### PAN-COVID

Pre-specified sample size estimation was not carried out given that the aim of this observational study was to collate all consecutive eligible case outcomes in participating centres through 18 months from the start of data collection. However, an approximate guide to the width of the 95% confidence intervals that can be achieved for proportions using historical data from the UK is given below.

The estimated representative pre-COVID pandemic incidences of miscarriage, SGA and stillbirth in the UK were 30%, 10%, 0.2% respectively. The prior assumptions of expected outcome proportions during the COVID pandemic were 40%, 15% and 0.4% respectively. A sample size of 500 would allow the estimation of the width of 95% confidence intervals for the proportion of miscarriage as 40 ± 4.2%, for SGA 15 ± 2.9% and for stillbirth 0.4 ± 0.3%. These figures are meant only as a guide and reference proportions may change over this period. These figures were based on proportions within the UK only; reference proportions vary by country but are of a similar order to those in the UK. Appropriate quantitative analyses were conducted by a dedicated study statistician. The PAN-COVID dataset was defined as those data records with delivery recorded up to and including 25/7/20.

**AAP SONPM National Registry** was conceived as an observational study and no a priori calculation of sample size was performed. The AAP SONPM registry contained information for all maternal/infant dyads entered as of 8/8/2020.

Gestational age at birth was calculated from expected due date (EDD) and the date of delivery recorded (the date of last menstrual period was used where EDD was unavailable). Calculated GA for inclusion, categorisation and birth-weight z-scores was limited as per the method in Fenton et. al^12^. Birthweight z-scores were gestational age- and gender-adjusted and limited to be within +/-4 standard deviations. Missing data for baby gender in PANCOVID restricted available data for z-scores such that the final data available was n=1423.Descriptive statistics are presented as numbers, percentages, and means with standard deviations as appropriate. The demography and outcomes between the two registries were compared using 95% confidence intervals for differences.

## Results

From January 1^st^ 2020 to July 24^th^ 2020, 1606 women were recruited to the PAN-COVID study; from April 1st 2020 to August 8^th^ 2020, 2398 women were recruited to the AAP SONPM registry. Symptomatic women with confirmed or putative COVID-19 and asymptomatic women who had positive swab tests for SARS-CoV-2 are described as *PAN-COVID all inclusions*; and those who had SARS-CoV-2 swab positive tests are described as *PAN-COVID* confirmed infection.

For PAN-COVID there were 1606 women in the data set at data censoring on 25/7/20, pregnancy outcomes were available for 1601 (99.7%), livebirths occurred in 1570 (98.1%), miscarriage in 23 (1.4%) and stillbirth/IUD in 8 women. Total livebirths of 1593 resulted from 1548 singleton, 21 twin, and 1 triplet gestation. Outcomes relating to the babies were available for 1454/1606 (90.5%) and for the purposes of calculating birthweight z-scores. Birthweight was available for n=1572. In the AAP SONPM registry, there were 2398 mothers with 2446 infants (including 58 twin births) at data censoring on 4/8/20. Outcome data were available for all mothers and infants.

Demographic details of the mothers are presented in Table 1 and key outcome measures are presented in Table 2. Women participating in PAN-COVID had a mean age of 32 years (SD 5.4) and AAP SONPM 28.6 years (SD 8.9). Body mass index (BMI) data were collected for PAN-COVID only. Maternal mean BMI was 27.8 (SD 6.4).

Ethnicity classifications were based on those relevant to the origins of the study and are not directly comparable. Women participating in PAN-COVID comprised 1066/1603 (66.5%) European/North American, 31 (1.9%) Middle Eastern, 18 (1.1%) North African, 67 (4.2%) were African south of the Sahara or Caribbean, 120 (7.5%) Indian subcontinent, 148 (9.2%) SE Asian, 13 (0.8%) South or Middle American and 140 (8.7%) ‘other’. Women participating in the AAP SONPM study comprised 905 (37%) white, 9 (0.4%) American Indian or Alaska Native American, 12 (0.5%) Native Hawaiian or Other Pacific Islander, 618 (25%) Black or African, 101 (4.1%) Asian, 722 (31%) Other, 35 (1.4%) Unknown or not recorded, and 7 (0.3%) Blank/Unanswered. For AAP SONPM there was almost an equal spread between Hispanic (n=1194) and non-Hispanic (n=1185) ethnicities (ethnicity was unknown or not reported in 19 women).

The PAN-COVID study collected data on pre-morbidities in pregnant women in both PAN-COVID all inclusions (n=1604) and PAN-COVID confirmed infection (n=651); the majority of the women in both cohorts had no pre-morbidities (63.7% and 61.9% respectively). Common pre-morbidities included gestational diabetes mellitus (131; 8.2% and 63; 9.7%), respiratory diseases such as asthma and COPD (96; 6% and 32; 4.9%), pregnancy induced hypertension (59; 3.7% and 30; 4.6%) and chronic hypertension (28; 1.7% and 15; 2.3%). The PAN-COVID study reported on smoking status of the mothers. Among all women (n=1600) and women with confirmed infection (n=647), 88 (5.5%) and 25 (3.9%) continued smoking, 200 (12.5%) and 51 (7.9%) stopped smoking before becoming pregnant, and 81 (5.1%) and 25 (3.9%) stopped smoking after conception, respectively.

Symptomatology of the mothers at presentation in the two studies are reported in Table 1. The main outcomes of the PAN-COVID and the AAP SONPM studies are presented in Table 2.

### Maternal mortality

Maternal death was reported in 8/1605 (0.50%) of PAN-COVID all inclusions, in 3/651 (0.46%) of PAN-COVID confirmed infections, and in 5/2398 (0.21%) of AAP SONPM registrants.

### Delivery

Vaginal delivery occurred in 880/1593 (55.2%) of PAN-COVID all inclusions, 334/641 (52.1%) of PAN-COVID confirmed infections and 1511/2398 (61.8%) of AAP SONPM participants. Pre-term delivery (< 37 weeks’ gestation) occurred in 190/1578 (12.0%) of PAN-COVID All inclusions cohort, in 103/635 (16.2%) of PAN-COVID Confirmed cohort and in 396/2398 (16.5%) of the AAP SONPM cohort (*Figure 1*). The majority of the preterm deliveries occurred late preterm (32 to 36 weeks’ gestational age). The rates of spontaneous (spontaneous labour or emergency Caesarean) preterm delivery in PAN-COVID and AAP SONPM were 114/190 (60.0%) and 173/396 (43.7%), respectively. Further details of the comparison of the main outcomes between PAN-COVID and AAP SONPM inclusions are presented in Table 3.

### Birthweight

The proportion of small for gestational age (SGA) infants (<10^th^ centile for gestational age birthweight) was 8.2% in the PAN-COVID all inclusions, 9.7% in the PAN-COVID confirmed infection and 9.6% in AAP SONPM (*Figure 2*). The mean birth weight z-score of AAP SONPM inclusions was significantly lower than PAN-COVID all inclusions cohort with mean z- = score (percentage point difference 0.14; CI 0.08 to 0.21); but there was no difference noted between PAN-COVID confirmed and the AAP SONPM cohorts.

### Perinatal mortality

Early neonatal death occurred in 3/1454 (0.2%) of PAN-COVID all inclusions, in 2/628 (0.3%) of PAN-COVID confirmed infection and in 8/2431, (0.3%) infants in the AAP SONPM. There were no differences between intra-uterine death and stillbirth which affected 8/1601 (0.5%) of the PAN-COVID all inclusions, 4/647 (0.6%) PAN-COVID confirmed infection and 15/2446 (0.65%) AAP SONPM (Table 3).

### Neonatal SARS-COV-2 infection

In AAP SONPM, neonatal screening was carried out in 2134/2431 (88%) of livebirths and in 1521593 (9.6%) of neonates in PAN-COVID all inclusions. Overall neonatal swab-positive for SARS-CoV-2 was seen in 44/2134 (2.1%) of the AAP SONPM, 15/1578 (0.8%) in the PAN-COVID all inclusions and 13/637 (2.2%) in the PAN-COVID confirmed infection.

## Discussion

Maternal deaths related to suspected or confirmed SARS-CoV-2 infection were uncommon in both the PAN-COVID and AAP SONPM studies. Maternal death before discharge from hospital affected a higher proportion compared to rates reported in UK population surveillance studies (9.8 women per 100,000 maternities)^13^. This is likely to be due to the low proportion of women in pregnancy with infection who were diagnosed and eligible for inclusion. By 1st July, 2020, 250,000 cases had been diagnosed in England and it was estimated that there had been 3.4 million infections i.e. less than 10% of infections were known^16,17^. When compared to the estimated infection fatality ratio in women and men aged 15-44 of 0.03% in the UK REACT 2 study^17^, the assumption that 10% of infections in women in pregnancy were diagnosed as cases would equate our reported mortality ratio to being closer to the expected infection fatality ratio.

On the other hand, the high perinatal maternal mortality rate in the AAP SONPM cohort of 167 per 100,000, compared to a pre-COVID rate of 17.3 per 100,000 through 1 year after birth, cannot be attributed to under inclusion of women with asymptomatic SARS-CoV-2 infection. Early in the pandemic, most US centres implemented universal COVID-19 testing of all pregnant women admitted to the labour and delivery unit. In both registries, where cause of death was known, all maternal deaths were associated with SARS-CoV-2 infection and hence each represents an additional mortality above the expected baseline rate.

Neither registry reported any neonatal death attributable to SARS-CoV-2. The proportion of pregnancies affected by early neonatal death (ENND) was no higher than would be expected from England and Wales ONS data (0.2%)^17^ or the USA from CDC data (0.38%) ^18^. An increase in prematurity might be expected to lead to an increase in ENND, however most preterm babies were born between 32 to 36 weeks’ gestation when risk for ENND is low.

In both studies, suspected or confirmed SARS-CoV-2 infection resulted in fewer than 10% of babies born small for gestational age (SGA, defined as birthweight <10^th^ centile for gestation) and did not change the expected distribution of birth weight z-scores. The proportion of pregnancies resulting in stillbirth, 1 in 200, is comparable to that reported in a UK population surveillance study (5.64 per 1000 total births)^19^, slightly greater than provisional Office of National Statistics (ONS) data for January to September 2020 (0.39%)^20^, and comparable to USA National Vital Statistics System data (611.7 per 100,000 live births) ^21^. This is reassuring because case reports of pregnant women with MERS or SARS infection suggested increased rates of stillbirth and fetal growth restriction^4^.

Another finding common to both registries was a high proportion of women with pre-term delivery, 12% in PAN-COVID and 16.5% in AAP SONPM. For PAN-COVID, this was 62% higher than expected for England and Wales based on ONS data for January-September 2020 (7.5%) ^20^ and for AAP SONPM 65% higher than expected based on US National Vital Statistics Reports for 2018 (10%)^22^. Pregnancies in which delivery occurred due to spontaneous pre-term labour or emergency Caesarean section accounted for 60% of pre-term deliveries in PAN-COVID and 40% in the AAP SONPM registries. There is no clear explanation for this because preterm birth was associated with both spontaneous and indicated deliveries. Nevertheless, a high proportion of pre-term deliveries may have been due to physician concern about adverse effects of SARS-CoV-2 infection on maternal or fetal condition.

Case series have reported low rates of perinatal transmission of SARS-CoV-2 infection ^10,23,24^. The proportion of positive neonatal tests for SARS-CoV-2 was approximately 9% in PAN-COVID all inclusions and 10% in PAN-COVID confirmed infection. A lower rate of 2% was found in the AAP SONPM registry. The difference in proportion reflects the near-universal testing strategy in AAP SONPM (2134/2431 (87%) neonates tested) and selective testing in PAN-COVID (152/1578 (9.4%) neonates tested).

The proportions of women who delivered by Caesarean section in PAN-COVID all inclusions and AAP SONPM were significantly higher than those reported by the OECD for the UK (27.4%) and the US (32%)^26^. Although reports from China from the beginning of the pandemic suggested an increase in the rate of Caesarean section deliveries,^24,27,28^ the caregivers planned Caesarean section whenever possible to minimize the risk of infection to the clinical team and infant.

### Strengths and weaknesses

This combined PAN-COVID and AAP SONPM report comprises the largest individual patient dataset of maternal and neonatal outcomes among women with suspected or confirmed SARS-CoV-2 infection. Standardised, online data-reporting with follow-up for missing data ensured a high proportion of complete data. Although there are some differences in populations (e.g., mean maternal age and race/ethnicity distributions), demographic and outcome data from these two registries are otherwise comparable and support the generalisability of the findings.

Both studies report more frequent preterm delivery than expected from contemporaneous and historical UK and US national data; the proportions for both outcomes are comparable between the studies despite the different healthcare settings.

In PAN-COVID, the higher than expected proportion of women included who died can be explained in part by under-ascertainment of pregnant women with SARS-CoV-2 who had mild or asymptomatic infection.

Data were collected from a range of healthcare settings, but the majority of participants originated from the US and UK with 11.1% of inclusions from PAN-COVID centres in Italy, China, Greece, Indonesia and India. This was a centre-based registry, centres with little exposure to COVID-19 or those which were overwhelmed during the pandemic may have been less motivated or able to participate.

This study does not allow conclusions to be drawn on the underlying factors relating to either the disease process, nor provision and utilisation of maternity services to evaluate the wider factors which influenced pregnancy, perinatal and neonatal outcomes^11^.

In PAN-COVID, it was difficult to associate SARS-CoV-2 infection directly with miscarriage. Patient inclusion occurred during the height of the pandemic, when many early-pregnancy units were closed in the UK. It is plausible that a higher proportion of women than usual may have miscarried without seeking help from a healthcare professional.

### Policy-makers and healthcare providers

Maternal death was uncommon, but a higher proportion of women participating in these studies died in comparison to contemporaneous and historical national rates of maternal mortality in the UK and US. In PAN-COVID, this was due in part to under-ascertainment of women with SARS-CoV-2 infection in pregnancy who had mild or asymptomatic infection.

It is reassuring that SARS-COV-2 infection does not appear to change the expected birth weight distribution or to increase the rate of stillbirth. The proportion of women with pre-term birth, as reported in other cohorts and meta-analyses ^25,29,30^, is higher than expected in comparison with national contemporaneous and historical data, but may have been influenced by provider decision to deliver early to prevent potential maternal or infant morbidity.

These data support public health guidance issued by numerous countries that advises pregnant women to exercise effective measures to reduce their risk of infection. Furthermore, these data support priority vaccination of women who plan to become pregnant to reduce the likelihood of preterm delivery and maternal mortality. Such guidance could be incorporated into pre-conception recommendations by national clinical advisory bodies.

The likelihood of and risk factors for true perinatal transmission to the neonate remain ill-defined and poorly understood. Recommendations for maternal and infant testing vary widely across countries and practices differ within countries. National and international healthcare bodies should develop recommendations for the timing and modality of testing of neonates born to pregnant women with SARS-COV-2 infection to facilitate consistent and meaningful evaluation of vertical transmission.

### Future work

Both studies continue to offer participation to all pregnant women who test positive for SARS-CoV-2 infection to March 2021. Future linkage to maternal and infant records will allow greater understanding of the longer-term outcomes for mothers and infants with the hope that mothers and infants at highest risk for serious complications of SARS-CoV-2 infection can be identified and appropriately counselled and treated.

## Supporting information

STROBE checklist

PAN COVID UK investigators

PAN COVID global investigators

AAP SONPM investigators

PAN COVID data fields

AAP SONPM data dictionary

## Data Availability

Anonymised data will be made available via the HDRUK portal after study completion in September 2021.

## Acknowledgements

PAN-COVID was funded by the UK National Institute for Health Research and supported by UK Clinical Research Network, Dr Nigel Simpson and the Urgent Public Health committee.

EM was funded by an NIHR Academic Clinical Lecturer award.

The AAP SONPM was funded by the University of Florida College of Medicine Division of Neonatology and by the in-kind contributions of participating centres.

We are grateful to Professor Helen Ward, Imperial College, for her comments on this manuscript.

The Imperial NIHR Biomedical Research Centre.

PAN-COVID study team: Rebecca Milton, Nigel Kirby, Matthew Robinson-Burt, Christopher Lloyd, Kim Munnery, Karina Aashamar

## References

1. Wong, SF et al. Pregnancy and perinatal outcomes of women with severe acute respiratory syndrome. Am. J. Obstet. Gynecol. 191, 292–297 (2004).

2. Li, A. & Ng, PC Severe acute respiratory syndrome (SARS) in neonates and children. Arch. Dis. Child. Fetal Neonatal Ed. 90, 461–465 (2005).

3. Shek, CC et al. Infants born to mothers with severe acute respiratory syndrome. Pediatrics 112, (2003).

4. Mullins, E, Evans, D, Viner, RM, O’Brien, P & Morris, E Coronavirus in pregnancy and delivery: rapid review. Ultrasound Obstet. Gynecol. 55, 586–592 (2020).

5. Lam, CM, Wong, F, Leung, N & Chow, M A case-controlled study comparing clinical course and outcomes of pregnant and non-pregnant women with severe acute respiratory syndrome. 111, 771–774 (2004).

6. Women with suspected COVID-19 or confirmed SARS-CoV-2 infection in pregnancy not admitted Confirmed SARS-CoV-2 infection Symptoms,. 19.

7. Pierce, M, Kurinczuk, JJ, Spark, P, Brocklehurst, P & Knight, M Perinatal outcomes after maternal 2009/H1N1 infection: National cohort study. BMJ 342, 1–8 (2011).

8. Liu, H et al. Why are pregnant women susceptible to viral infection: an immunological viewpoint? J. Reprod. Immunol. (2020) doi:https://doi.org/10.1016/j.jri.2020.103122.

9. Knight, M et al. Characteristics and outcomes of pregnant women hospitalised with confirmed SARS-CoV-2 infection in the UK: a national cohort study using the UK Obstetric Surveillance System (UKOSS) The UK Obstetric Surveillance System SARS-CoV-2 Infection in Pregnancy Co. (2020) doi:https://doi.org/10.1101/2020.05.08.20089268.

10. Vivanti, AJ et al. Transplacental transmission of SARS-CoV-2 infection. Nat. Commun. (2020) doi:10.1038/s41467-020-17436-6.

11. Ashish, KC et al. Effect of the COVID-19 pandemic response on intrapartum care, stillbirth, and neonatal mortality outcomes in Nepal: a prospective observational study. Lancet Glob. Heal. (2020) doi:10.1016/S2214-109X(20)30345-4.

12. Fenton, T. & Kim, JH A systematic review and meta-analysis to revise the Fenton growth chart for preterm infants. BMC Pediatr. 13, (2013).

13. MBRRACE. Saving Lives, Improving Mothers’ CareLessons learned to inform maternity care from theUK and Ireland Confidential Enquiries into MaternalDeaths and Morbidity 2014–16. Maternal, Newborn andInfant Clinical Outcome Review Programme (2018).

14. Chinn, JJ et al. Maternal mortality in the United States: research gaps, opportunities, and priorities. Am. J. Obstet. Gynecol. 223, 486–492.e6 (2020).

15. CDC Centers for Disease Control and Prevention. Pregnancy mortality surveillance system. 2016. https://www.cdc.gov/reproductivehealth/maternal-mortality/pregnancy-mortality-surveillance-system.htm.

16. Ward, H et al. Antibody prevalence for SARS-CoV-2 following the peak of the pandemic in EnglandllJ: REACT2 study in 100, 000 adults Corresponding authors. MedRxiv 1–20 (2020).

17. ONS Infant mortality (birth cohort) tables in England and Wales. https://www.ons.gov.uk/peoplepopulationandcommunity/birthsdeathsandmarriages/deaths/datasets/infantmortalitybirthcohorttablesinenglandandwales (2017).

18. Ely, D. & Driscoll, A. K. Infant mortality in the United States, 2018: Data from the period linked birth/infant death file. Natl. Vital Stat. Reports 69, 1–17 (2020).

19. Draper, E. et al. MBRRACE-UK Perinatal Mortality Surveillance Report, UK Perinatal Deaths for Births from January to December 2016. Leicester: The Infant Mortality and Morbidity Studies, Department of Health Sciences, University of Leicester. (2018).

20. ONS Provisional births in England and Wales: 2020. Births, deaths and marriages (2020).

21. Hoyert, D. & Gregory, EC W. Cause of Fetal Death: Data From the Fetal Death Report, 2014. Natl. Vital Stat. Rep. 65, 1–25 (2016).

22. Martin, JA, Hamilton, BE, Osterman, MJK & Driscoll, AK Births: Final data for 2018. Natl. Vital Stat. Reports 68, 1980–2018 (2019).

23. Yang, P et al. Clinical characteristics and risk assessment of newborns born to mothers with COVID-19. J. Clin. Virol. (2020) doi:10.1016/j.jcv.2020.104356.

24. Liu, W et al. Clinical characteristics of 19 neonates born to mothers with COVID-19. Front. Med. (2020) doi:10.1007/s11684-020-0772-y.

25. Knight, M et al. Characteristics and outcomes of pregnant women admitted to hospital with confirmed SARS-CoV-2 infection in UK: National population based cohort study. BMJ 369, (2020).

26. OECD iLibrary. Health at a Glance 2019: OECD Indicators. Caesarean sections https://www.oecd-ilibrary.org/sites/fa1f7281-en/index.html?itemId=/content/component/fa1f7281-en (2019).

27. Yu, N et al. Clinical features and obstetric and neonatal outcomes of pregnant patients with COVID-19 in Wuhan, China: a retrospective, single-centre, descriptive study. Lancet Infect. Dis. 20, (2020).

28. Li, N et al. Maternal and neonatal outcomes of pregnant women with COVID-19 pneumonia: a case-control study. Clin. Infect. Dis. (2020) doi:10.1093/cid/ciaa352.

29. Allotey, J et al. Clinical manifestations, risk factors, and maternal and perinatal outcomes of coronavirus disease 2019 in pregnancy: Living systematic review and meta-analysis. BMJ 370, (2020).

30. Flaherman, VJ et al. Infant Outcomes Following Maternal Infection With Severe Acute Respiratory Syndrome Coronavirus 2 (SARS-CoV-2): First Report From the Pregnancy Coronavirus Outcomes Registry (PRIORITY) Study. Clin. Infect. Dis. 2, 1–4 (2020).

