## Supplementary material for "Pregnancy and neonatal outcomes of COVID-19 – co-reporting of common outcomes from the PAN-COVID and AAP SONPM registry": Figure 1

*Figure 1. Gestational age at birth for mothers recruited to PAN-COVID all inclusions, PAN-COVID confirmed infection and AAP-SONPM registries*

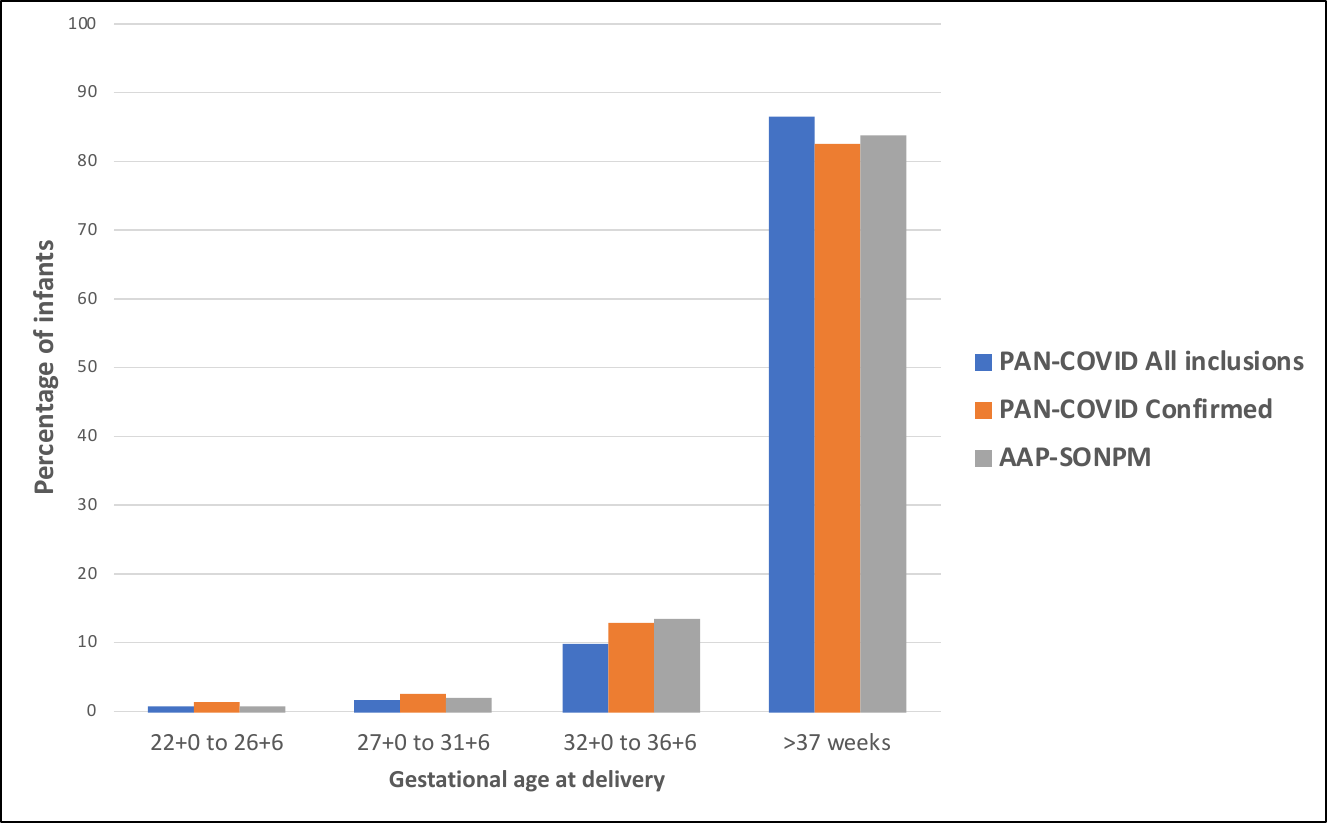
