## Supplementary material for "Pregnancy and neonatal outcomes of COVID-19 – co-reporting of common outcomes from the PAN-COVID and AAP SONPM registry": Figure 2

*Figure 2. Birth weight percentiles of infants born of mothers in PAN-COVID all inclusions, PAN-COVID confirmed infection and AAP-SONPM studies*

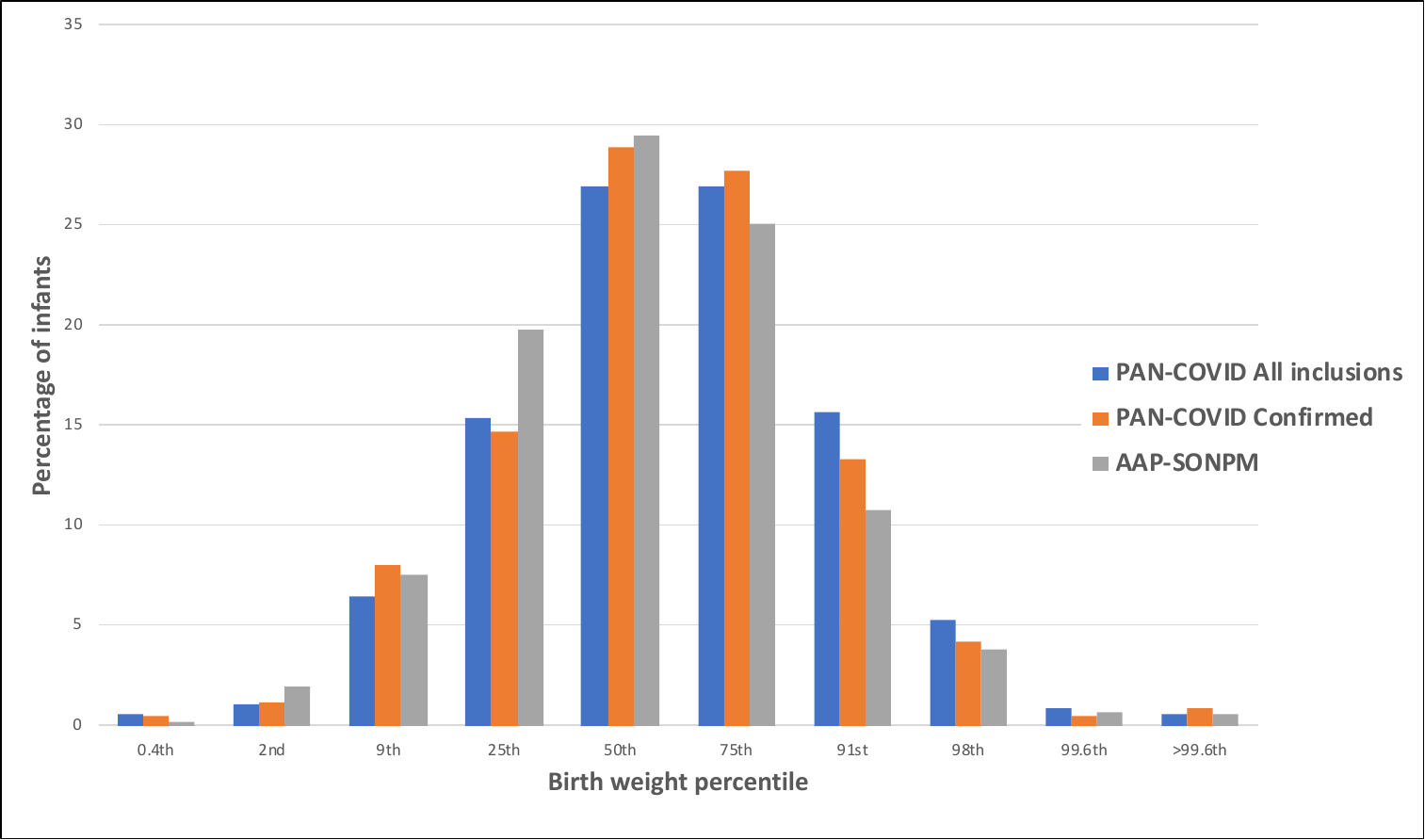
