## Supplementary material for "Pregnancy and neonatal outcomes of COVID-19 – co-reporting of common outcomes from the PAN-COVID and AAP SONPM registry": Table 1

**Table 1.** Maternal demographics and symptomatology

|  | N | **PAN-COVID (all inclusions)** | N | **PAN-COVID (confirmed infection)** | N |  | **AAP SOPNM** |
| --- | --- | --- | --- | --- | --- | --- | --- |
|  |  | Mean (SD) |  | Mean (SD) |  |  | Mean (SD) |
| Age at registration (years) | 1606 | 32.0 (5.4) | 651 | 31.8 (5.5) | 2398 |  | 28.6 (8.9) |
| BMI (kg/m^2^) | 1585 | 27.8 (6.4) | 6536 | 28.2 (6.2) | - |  | - |
|  |  | **n (%)** |  | **n (%)** |  |  | **n (%)** |
| **Maternal symptoms at presentation (%yes)** | 1216 |  | 349 |  |  |  | 2398 |
| Asymptomatic |  |  |  |  |  |  | 1820 (75.9%) |
| Fever |  | 588 (48.4%) |  | 134 (38.4%) |  | Fever | 195 (8.1%) |
| New, persistent cough |  | 687 (56.5%) |  | 130 (37.2%) |  | URTI | 337 (14.1%) |
| Shortness of breath |  | 343 (28.2%) |  | 77 (22.1%) |  | LRTI | 143 (6.0%) |
| Chest pain |  | 121 (10.0%) |  | 20 (5.7%) |  |  |  |
| Anosmia |  | 229 (18.8%) |  | 36 (10.3%) |  | Anosmia/ageusia | 75 (3.1%) |
| Hoarse voice |  | 101 (8.3%) |  | 8 (2.3%) |  |  |  |
| Myalgia |  | 203 (16.7%) |  | 34 (9.7%) |  | Myalgia and fatigue | 118 (4.9%) |
| Fatigue |  | 381 (31.3%) |  | 49 (14.0%) |  |  |  |
| Diarrhoea |  | 73 (6.0%) |  | 22 (6.3%) |  | GI symptoms (Diarrhoea, vomiting, nausea) | 63 (2.6%) |
| Loss of appetite |  | 134 (11.0%) |  | 24 (6.9%) |  |  |  |
| Abdominal pain |  | 34 (2.8%) |  | 6 (1.7%) |  |  |  |
| Delirium |  | 14 (1.2%) |  | 1 (0.3%) |  |  |  |
| None of the above |  | 159 (13.1%) |  | 134 (38.4%) |  | Other/nothing selected | 99 (4.1%) |
| Ethnicity | 1603 |  | 648 |  | 2398 |  |  |
| European / North American |  | 1066 (66.5%) |  | 319 (49.2%) |  |  | 905 (37.7%) |
| Middle East |  | 31 (1.9%) |  | 11 (1.7%) |  |  |  |
| Northern Africa |  | 18 (1.1%) |  | 9 (1.4%) |  |  |  |
| Africa South of Sahara / Caribbean |  | 67 (4.2%) |  | 38 (5.9%) |  |  | 618 (25.8%) |
| Indian subcontinent |  | 120 (7.5%) |  | 79 (12.2%) |  |  | 101 (4.2%) |
| SE Asia |  | 148 (9.2%) |  | 108 (16.7%) |  |  |  |
| South - Middle America |  | 13 (0.8%) |  | 11 (1.7%) |  |  |  |
| Other |  | 140 (8.7%) |  | 73 (11.3%) |  |  | 722 (32.2%) |
