## Supplementary material for "Pregnancy and neonatal outcomes of COVID-19 – co-reporting of common outcomes from the PAN-COVID and AAP SONPM registry": Table 2

**Table 2.** Maternal and neonatal outcomes of confirmed or suspected SARs-CoV-2 infection in pregnancy

| **Outcomes** | **PAN-COVID**  **(all inclusions)** | **PAN-COVID**  **(confirmed infection)** | **AAP SONPM** |
| --- | --- | --- | --- |
|  | **N (%)** | **N (%)** | **N (%)** |
| Maternal death | 8/1605 (0.5%) | 3/651 (0.5%) | 5/2398 (0.21%) |
| Early neonatal death | 3/1454 (0.2%) | 2/628 (0.3%) | 8/2446 (0.3%) |
| Pregnancy outcomes | N=1601* | N=647* | N = 2398* |
| Liveborn | 1570 (98.1%) | 631 (97.5%) | 2429 (99.3 %) |
| Miscarriage | 23 (1.4%) | 12 (1.9%) | 5 (0.2%) |
| Intra-uterine death/stillbirth (>22+0 weeks Gestation) | 8 (0.5%) | 4 (0.6%) | 10(0.4%) |
| Mode of delivery (all births) | N=1593 | N=641 | N=2398 |
| Vaginal | 880 (55.2%) | 334 (52.1%) | 1511 (61.8%) |
| C section | 713 (44.8%) | 307 (47.9%) | 933 (38.2%) |
| Pre-term delivery  (23+0 to 36+6 weeks) | N=190/1578 (12.0%)* | N= 103/635 (16.2%)* | N=396/2398 (16.5%)* |
| Gestational age at birth  (22+0 to 45 weeks) all births | N=1561* | N=628* | N=2416* |
| 22+0 to 26+6 weeks | 10 (0.6%) | 8 (1.3%) | 12 (0.57%) |
| 27+0 to 31+6 weeks | 25 (1.6%) | 16 (2.5%) | 49 (2.1%) |
| 32+0 to 36+6 weeks | 158 (10.1%) | 82 (13.0%) | 330 (13.5%) |
| 37+0 to 45+0 weeks | 1369 (87.6%) | 523 (83.1%) | 2050 (83.8%) |
| Gestational age at birth  (22+0 to 45 weeks) live births | N=1555* | N=622* | N=2431 |
| 22+0 to 26+6 weeks | 7 (0.5%) | 5 (0.8%) |  |
| Birth weight outcomes (all singletons and first-born twin with GA between 22 and 49 weeks and z-score between +/- 4) | N=1423* | N=577* | N=2421* |
| Percentiles | n (%) | n (%) | n (%) |
| 0.4^th^ | 9 (0.6%) | 3 (0.5%) | 4 (0.2%) |
| 2^nd^ | 16 (1.1%) | 7 (1.2%) | 46 (1.9%) |
| 9^th^ | 92 (6.5%) | 46 (8.0%) | 182 (7.5%) |
| 25^th^ | 219 (15.4%) | 85 (14.7%) | 480 (19.8%) |
| 50^th^ | 383 (26.9%) | 167 (28.9%) | 714 (29.5%) |
| 75^th^ | 383 (26.9%) | 160 (27.7%) | 607 (25.1%) |
| 91^st^ | 224 (15.7%) | 77 (13.3%) | 262 (10.8%) |
| 98^th^ | 75 (5.3%) | 24 (4.2%) | 93 (3.8%) |
| 99.6^th^ | 13 (0.9%) | 3 (0.5%) | 18 (0.7%) |
| >99.6^th^ | 9 (0.6%) | 5 (0.9%) | 15 (0.6%) |
| Birth weight z-scores | Mean (SD) | Mean (SD) | Mean (SD) |
| All singleton and first-born twin** | z -0.03 (0.95) | z -0.10 (0.94) | z -0.18 (0.94) |
| Singletons only (n=1391) | z -0.03 (0.91) | z -0.11 (0.89) |  |
| Neonatal SARS-CoV-2 testing | N (%) | N (%) | N (%) |
| Neonatal COVID-19 or SARS-CoV-2 swab positive (% of those tested) | 14/152 (9.2%) | 13/131 (9.9%) | 44/2134 (2.1%) |
| Neonatal COVID-19 or SARS-CoV-2 swab positive (% of all deliveries) | 14/1578 (0.8%) *** | 13/647 (2%)*** | 44/2444 (1.8%) |

* Available data presented.

**For PAN-COVID 1^st^ born twin birthweights were collected, for AAP SONPM all babies’ birthweights were included

*** not all neonates tested, presented for comparison with AAP SONPM
