## Supplementary material for "Pregnancy and neonatal outcomes of COVID-19 – co-reporting of common outcomes from the PAN-COVID and AAP SONPM registry": Table 3

*Table 3: Percentage point differences between PAN-COVID all inclusions vs AAP SONPM, and PAN-COVID confirmed infection vs AAP SONPM.*

|  | PANCOVID all inclusions vs AAP SONPM | PANCOVID confirmed infection vs AAP SONPM |
| --- | --- | --- |
|  | Percentage point difference and 95% CI | Percentage point difference and 95% CI |
| Preterm delivery  (23+0 to 36+6 weeks) | -4.2 (-6.3 to -2.0) | 0.02 (-3.1 to 3.4) |
| Inter-uterine death/stillbirth  (>22+6 weeks Gestation) | -0.15 (-0.63 to 0.39) | -0.04 (-0.59 to 0.96) |
| Early neonatal death | 0.21 (0 to 0.6) | 0.34 (0.05 to 1.22) |
