## Supplementary material for "Pregnancy and neonatal outcomes of COVID-19 – co-reporting of common outcomes from the PAN-COVID and AAP SONPM registry": Table 4

Table 4 Summary characteristics of PAN-COVID and AAP SONPM registries

|  | **PAN-COVID** | **AAP SONPM** |
| --- | --- | --- |
| **Study period** | 01/01/20-25/07/20 | 01/04/2020-08/08/20 |
| **Population(s)** | UK & global | USA |
| **Inclusion criteria** | Suspected COVID-19 or confirmed SARS-CoV-2 at any stage in pregnancy | Positive SARS-CoV-2 test from 14 days before to 3 days after delivery |
