## Supplementary material for "Pregnancy and neonatal outcomes of COVID-19 – co-reporting of common outcomes from the PAN-COVID and AAP SONPM registry": PAN COVID UK investigators

**PAN-COVID project partners** | **(planning funding application, study conduct)**

| | |
|---|---|
| **PAN-COVID project partners** | **(planning funding application, study conduct)** |
| Dr Mandish Dhanjal | Imperial College Healthcare NHS Trust |
| Professor Tg Teoh | Imperial College Healthcare NHS Trust, Imperial College London |
| Dr Alison Wright | Royal Free NHS Foundation Trust, London |
| Professor Liona Poon | Chinese University of Hong Kong |
| Alison Perry | Manager, Women's Health Research Centre, Imperial College London |
| Dr Caroline Shaw | Clincial Lecturer, Imperial College London |
| Professor Dirk Timmerman | KU Leuven, Belgium |
| Professor Neil Ferguson | Imperial College London |
| Professor Neena Modi | Imperial College London |

**PAN-COVID Investigators**

| Trust Name | Site | Principal Investigator | RM / Coordinator |
|---|---|---|---|
| Airedale NHS Foundation Trust | | Soum Nallapeta | emma dooks |
| Aneurin Bevan University Health Board | Royal Gwent Hospital | Emma Mills | Tracy James |
| Ashford and St Peter's Hospitals NHS Foundation Trust | St Peter's Hospital | Beth Peers | Hayley Tarft |
| Barnsley Hospital NHS Foundation Trust | | Sarah Stables | Allison Daniels |
| Barts Health NHS Trust | Royal London Hospital | Dr Stamatina Iliodromiti | Megan Parrott |
| Barts Health NHS Trust | Newham Hospital | | Tabitha Newman |
| Barts Health NHS Trust | Whipps Cross Hospital | | Amy Thomas |
| Betsi Cadwaladr University Health Board | Wrexham Maelor | Maggie Armstrong | SARAH DAVIES |
| Betsi Cadwaladr University Health Board | Ysbyty Gwynedd | Hilary Owen | |
| Betsi Cadwaladr University Health Board | Glan Clwyd | | MEL HOLLINS |
| Birmingham Women's and Children's NHS Foundation | Birmingham Women's Hospital | Shanteela Mccooty | Amy Woodhead |
| Black Country Healthcare NHS Foundation trust | Dorothy Pattison Hospital | Dr Anila Asghar | Florentina Takacs |
| Blackpool Teaching Hospitals NHS Foundation Trust | Blackpool Victoria Hospital | Dr Eric Mutema | Emma Stoddard |
| Bolton NHS Foundation Trust | Royal Bolton | Emma Tanton | Kat Rhead |
| Bradford Teaching Hospitals NHS Foundation Trust | | Jen Syson | Jenny Eedle |
| Buckinghamshire Healthcare NHS Trust | Stoke Mandeville | Danielle Thornton | Lisa Frankland |
| Calderdale And Huddersfield NHS Foundation Trust | Calderdale Royal Hospital | Julie Goddard | Marie Home |
| Calderdale And Huddersfield NHS Foundation Trust | Huddersfield Royal Infirmary | Julie Goddard | Kelly Holroyd |
| Cambridge University Hospitals NHS Foundation Trust | Rosie Hospital | Elena Romero | Amy Sutton-Cole |
| Cardiff and Vale UHB | UHW | Maryanne Bray | Vikki Keeping |
| Chelsea and Westminster Hospital NHS Foundation Trus | Chelsea and Westminster Hospital | Miriam Bourke | Natasha Singh |
| Chelsea and Westminster Hospital NHS Foundation Trus | West Middlesex University Hospital | Lauren Trepte | Amy West |
| Chesterfield Royal Hospital NHS Foundation Trust | Chesterfield Royal Hospital | Janet Cresswell | Mary Kelly-Baxter |

| | | | |
|---|---|---|---|
| Countess of Chester Hospital NHS Foundation Trust | | Dr Trevor Balling | Kerry Barker-Williams |
| County Durham and Darlington NHS Foundation Trust | Darlington Memorial Hospital | Vicki Atkinson | Jacqui Jennings |
| County Durham and Darlington NHS Foundation Trust | University Hospital of North Durham | | |
| Croydon University Hospital NHS Trust | Croydon University Hospital | Bini Ajay | Gerry Upson |
| Cwm Taf Morganwnwg University Health Board | Princess of Wales Hospital | Lavinia Margarit | Joelle Pike |
| Cwm Taf Morganwnwg University Health Board | Prince Charles Hospital | | Annabel Creeth |
| Dartford and Gravesham NHS Trust | Darent Valley Hospital | Samirah Toure | |
| Devon Partnership NHS Trust | Wonford Hospital | Laurie Windsor | Anna Grice |
| Dorset County Hospital NHS Foundation Trust | Dorset County Hospital | Donna Wixted | Heather Sellers |
| East and North Hertfordshire NHS Trust | Lister Hospital | Miss Rabia Zill-E-Huma | Sarah Johnson |
| East Kent Hospitals University NHS Foundation Trust | The William Harvey Hospital | Dr Vimal Vasu | James Rand |
| East Kent Hospitals University NHS Foundation Trust | Queen Elizabeth the Queen Mother Ho | Dr Zoe Woodward | Tracy Hazelton |
| East Lancashire Hospitals NHS Trust | Burnley General Hospital | Beverley Hammond | Laura Hoole |
| East Lancashire Hospitals NHS Trust | Royal Blackburn Teaching Hospital | | |
| East Suffolk and North Essex NHS Foundation Trust | Colchester Hospital | Wassim Hassan | Dr Sasha Taylor |
| East Suffolk and North Essex NHS Foundation Trust | Ipswich Hospital | Dr Ruta Gada | Dr Samantha Parlapalli |
| East Sussex Healthcare NHS Trust | Conquest Hospital | Nicky Mason | Gayle Clarke |
| East Sussex Healthcare NHS Trust | Eastbourne District General Hospital | Nicky Mason | |
| Epsom and St Helier University Hospitals NHS Trust | St Helier Hospital | ouise Emmet, Consultant Midw | Katherina Gross-Gibbs |
| Epsom and St Helier University Hospitals NHS Trust | Epsom Hospital | ouise Emmet, Consultant Midwif | |
| Frimley Health NHS Foundation Trust | Frimley Park | Lianne Chapman | Alex Edwards |
| Frimley Health NHS Foundation Trust | Wexham Park | Sarah Coxon | Catherine Smith |
| Gateshead Health NHS Foundation Trust | QEH Gateshead | Christine Moller-Christensen | Rachael Grant |
| George Eliot Hospital NHS Trust | | Shazia Jaleel | Tracy Truslove |
| Gloucestershire Hospitals NHS Foundation Trust | Gloucestershire Royal Hospital | Mrs Siân .C. Harrington | |
| Great Western Hospitals NHS Foundation Trust | Great Western Hospital | Ruth Davies | |
| Guy's and St Thomas' NHS Foundation Trust | St Thomas' Hospital | Dr Caroline Knight | Alice Lewin |
| Hampshire Hospital NHS Foundation Trust | Royal Hampshire County Hospital | Kirsty Revell | Ana Maria Arias |
| Hampshire Hospital NHS Foundation Trust | Basingstoke and North Hampshire Hosp | Avideah Nejad | Tracey Dunham |
| Harrogate And District NHS Foundation Trust | Harrogate District Hospital | Mrs Allison Amin | Louise Willis |
| Homerton University Hospital NHS Foundation Trust | Homerton University Hospital | Narendra Aladangady | Asha Mathew |
| Hull University Teaching Hospitals NHS Trust | | Leanne Sherris | Melony Bowdler-Hayes |

| | | | |
|---|---|---|---|
| Imperial College Healthcare NHS Trust | Queen Charlotte & Chelsea Hospital | Edward Mullins,<br>Roshni Mansfield | Alison Perry |
| Imperial College Healthcare NHS Trust | St Marys Hospital | | Jenny Goodier |
| Isle of Wight NHS Trust | St Mary’s Hospital | | Elinor Jenkins |
| James Paget University Hospitals NHS Foundation Trust | | Jamie-Louise Raven | Joanna Keable |
| King's College Hospital NHS Foundation Trust | Princess Royal University Hospital | Hayley Martin | Gillian Goodwin |
| King's College Hospital NHS Foundation Trust | Denmark Hill | Hayley Martin | Katherine Clark |
| Kingston Hospital NHS Foundation Trust | Kingston Hospital | Changing PI - no update yet | Kingston Maternity |
| Lancashire Teaching Hospitals NHS Foundation Trust | | Cheryl Wyatt | Julie Earnshaw |
| Leeds Teaching Hospitals NHS Trust | St James University Hospital | Kate Robinson | Jayne Wagstff |
| Leeds Teaching Hospitals NHS Trust | Leeds General Infirmary | Kate Robinson | |
| Lewisham and Greenwich NHS Trust | University Hospital Lewisham | Muglu Javaid | Chloe Saad |
| Lewisham and Greenwich NHS Trust | Queen Elizabeth Hospital Woolwich | Veerareddy Sukrutha | |
| Liverpool Women's NHS Foundation Trust | | Amy Mahdi | Siobhan Holt |
| London North West University Healthcare NHS Trust | Northwick Park Hospital | Anam Fayadh | |
| Maidstone and Tunbridge Wells NHS Trust | Tunbridge Wells Hospital | Louise Swaminathan | Philippa Hadlow |
| Maidstone and Tunbridge Wells NHS Trust | Maidstone Hospital | | |
| Manchester University NHS Foundation Trust | St Mary’s | Sam Ratcliffe | Meg Hyslop |
| Medway NHS Foundation Trust | Medway Maritime Hospital | Helen Gbinigie | Sarah-Jayne Ambler |
| Mid and South Essex Hospital Services NHS Trust | Broomfield Hospital | Ms Sameena Kausar | Sandeep Virdee |
| Mid and South Essex Hospital Services NHS Trust | Southend University Hospital | Andrea Harrington | Eugene Mphansi |
| Mid and South Essex Hospital Services NHS Trust / Basil | Basildon University Hospital | Donna Southam | Stacey Pepper |
| Mid Cheshire Hospitals NHS Foundation Trust | Leighton Hospital/SAA | Emily Lear | Caroline Dixon |
| Mid Yorkshire Hospitals NHS Trust | Pinderfields Hospital | Dr Rukhsana Kousar | Gail Castle |
| Mid Yorkshire Hospitals NHS Trust | Pontefract Hospital | Dr Rukhsana Kousar | |
| Mid Yorkshire Hospitals NHS Trust | Dewsbury District Hospital | Dr Rukhsana Kousar | |
| Milton Keynes University Hospital NHS Foundation Trust | Milton Keynes University Hospital | Joanna Mead | Edel Clare |
| NHS Grampian | Aberdeen Maternity Hospital | Dr Mairead Black | |
| NHS Grampian | Aberdeen Maternity Hospital | | Minimol Paulose |
| NHS Greater Glasgow & Clyde | Queen Elizabeth University Hospital | Isobel Crawford | Christine Campbell |
| NHS Greater Glasgow & Clyde | Princess Royal Maternity | | |
| NHS Greater Glasgow & Clyde | Royal Alexandra Hospital | | |
| NHS Greater Glasgow & Clyde | Glasgow Royal Infirmary | | |

| | | | |
|---|---|---|---|
| NHS Lothian | Royal Infirmary of Edinburgh, St John's I | Dr Alexandra Viner | |
| NHS Tayside | Ninewells Hospital | Dr Antony Nicoll | |
| NHS Tayside | Perth Royal Infirmary | Dr Antony Nicoll | |
| NHS Tayside | Arbroath Infirmary | Dr Antony Nicoll | |
| NHS Tayside | Montrose Infirmary | Dr Antony Nicoll | |
| Norfolk and Norwich University Hospitals NHS Foundati | Norfolk and Norwich University Hospita | Laura Harris | Louise Coke (Co PI) |
| North Bristol NHS Trust | Southmead Hospital | Nichola Bale | Mary Alvarez |
| North Cumbria Integrated Care NHS Foundation Trust | The Cumberland Infirmary | Mr Bilal Rather | Rachel Hardy |
| North Cumbria Integrated Care NHS Foundation Trust | West Cumberland Hospital | Mr Bilal Rather | |
| North Middlesex University Hospital NHS Trust | North Middlesex Hospital | Mrs Sandra Essien | Dr Abha Govind |
| North Tees and Hartlepool NHS Foundation Trust | North Tees | Sharon Gowans | Alex Ramshaw |
| North West Anglia NHS Foundation Trust | Peterborough City Hospital | Coralie Huson | Jodie Carpenter |
| North West Anglia NHS Foundation Trust | Hitchinbrooke Hospital | Coralie Huson | Kimberly Morries |
| Northumbria Healthcare NHS Foundation Trust | Northumbria healthcare NHS Foundatio | Katie Barker | Cath Ashbrook-Raby |
| Nottingham University Hospitals NHS Trust | Nottingham City Hospital | Jane Cantliffe | Harriet Anderson |
| Nottingham University Hospitals NHS Trust | Queens Medical Centre | Lesley Hodgen | |
| Oxford University Hospitals NHS Foundation Trust | John Radcliffe Hospital | Jude Mossop | Lisa Buck |
| Pennine Acute Hospitals NHS Trust | Royal Oldham Hospital | Rachel Newport | Rachel Newport |
| Pennine Acute Hospitals NHS Trust | Fairfield General Hospital | Rachel Newport | |
| Pennine Acute Hospitals NHS Trust | Rochdale Infirmary | Rachel Newport | |
| Pennine Acute Hospitals NHS Trust | North Manchester General Hospital | Rachel Newport | |
| Poole Hospital NHS Foundation Trust | St Mary’s Maternity - Poole | Susara Blunden RM | Stephanie Grigsby |
| Portsmouth Hospitals NHS Trust | Queen Alexandra Hospital | Zoe Garner | |
| Powys Teaching Health Board – Ystradgynlais | Ystradgynlais, Brecon, Llandrindod Well | Shelly Higgins | Liz Glyn-Jones |
| Royal Berkshire NHS Foundation Trust | | Fidelma Lee | Lianne Chapman |
| Royal Cornwall Hospitals NHS Trust | | Karen Watkins | Ali Dorning |
| Royal Devon and Exeter NHS Foundation Trust | | Jacqueline Tipper | Caroline Blake |
| Royal Free London NHS Foundation Trust | Royal Free Hospital | Michelle Anderson | Eleanor Pyart |
| Royal Free London NHS Foundation Trust | Barnet Hospital | | |
| Royal Surrey NHS Foundation Trust | Royal Surrey Hospital NHS Foundation T | Caroline Everden | Georgina Black |
| Royal United Hospitals Bath NHS Foundation Trust | RUH | Catherine Bressington | Sara Burnard |
| Salisbury NHS Foundation Trust | Salisbury District Hospital | Abby Rand | Holly Morgan |
| Sandwell and West Birmingham Hospitals NHS Trust | City Hospital Birmingham | Neil Shah | Lavinia Henry |

| | | | |
|---|---|---|---|
| Sandwell and West Birmingham Hospitals NHS Trust | Sandwell General Hospital | | |
| Sheffield Teaching Hospitals NHS Foundation Trust | Jessop Wing Hospital | Roobin Jokhi | general email |
| Sherwood Forest Hospitals NHS Foundation Trust | Kings Mill Hospital | Jyothi Rajeswary | Many Gill |
| Shrewsbury and Telford Hospital NHS Trust | Shrewsbury and Telford Hospital | Helen Millward | Tracie Kenny |
| Somerset NHS Foundation Trust | | Ami Mackay | Kirsty O'Brien |
| South Tees Hospitals NHS Foundation Trust | James Cook University Hospital | Aethele Khunda | Helen Harwood |
| South Tyneside and Sunderland NHS Foundation Trust | Sunderland Royal Hospital | nna Ahmed. Co-PI is Kim Hinsh | Gemma Parish |
| South Tyneside and Sunderland NHS Foundation Trust | South Tyneside District Hospital | nna Ahmed. Co-PI is Kim Hinshaw | |
| South Warwickshire NHS Foundation Trust | | Clare O'Brien | Kelly Jukes |
| Southern Health and Social Care Trust | Daisy Hill Hospital | Dr Gillian McKeown | Fiona Thompson |
| Southern Health and Social Care Trust | Craigavon Hospital | Dr Gillian McKeown | |
| Southport and Ormskirk Hospital NHS Trust | | Linda Bishop | ZENA HASLAM |
| St George's University Hospitals NHS Foundation Trust | St George's Hospital | Sophie Robinson | Danielle Hake |
| St Helens and Knowsley Hospital Services NHS Trust | Whiston Hospital | Sandra Greer | |
| St Helens and Knowsley Hospital Services NHS Trust | St Helens Hospital | Sandra Greer | |
| Stockport NHS Foundation Trust | Stepping Hill Hospital | Carrie Heal | Sara Bennett |
| Surrey and Sussex Healthcare NHS Trust | East Surrey Hospital | Mahalakshmi Gorti | Sarah Maher |
| Swansea Bay University Health Board | Singleton Hospital (all sites) | Sharon Jones | Eve Watkins |
| Tameside and Glossop Integrated Care NHS Foundation | Tameside Hospital | Millicent Anim-Somuah | Elena Ruding |
| NHS Lanarkshire | University Hospital Wishaw | Eleanor Jarvie | Denise Vigni |
| The Hillingdon Hospitals NHS Foundation Trust | Hilligdon Hospital | Laura Camarasa | Komal Lal |
| The Newcastle upon Tyne Hospitals NHS Foundation Tru | Royal Victoria Infirmary | Victoria Murtha | Fiona Yelnoorkar |
| The Newcastle upon Tyne Hospitals NHS Foundation Tru | Newcastle Hospitals | | Jill Riches |
| The Princess Alexandra Hospital NHS Trust | The Princess Alexandra Hospital | Mr Ling Wee | Nikki Staines |
| The Queen Elizabeth Hospital King's Lynn NHS Foundatic | Queen Elizabeth Hospital Kings Lynn | Mr Salman Kidwai | Zoe Coton |
| The Royal Wolverhampton NHS Trust | New Cross Hospital | Mr David Churchill | Laura Devison |
| Torbay and South Devon NHS Foundation Trust | Torbay Hospital | KAREN CLOHERTY | CATHERINE MARSHALL |
| United Lincolnshire Hospitals NHS Trust | Pilgrim Hospital Boston | Chris Flood | Kimberley Netherton |
| United Lincolnshire Hospitals NHS Trust | Lincoln County Hospital | | |
| University College London Hospitals NHS Foundation Tru | University Hospital London | Sarah Ekladios | Erin Lever |
| University Hospital Southampton NHS Foundation Trust | Princess Anne Hospital | Dr Alexandra Kermack | Sue Wellstead |
| University Hospital Southampton NHS Foundation Trust | University Hospital Southampton | | |

| | | | |
|---|---|---|---|
| University Hospitals Birmingham NHS Foundation Trust | Birmingham Heartlands Hospital | Mani Malarselvi | Lucy O'Leary |
| University Hospitals Birmingham NHS Foundation Trust | Good Hope Hospital | Vibha Giri | Rujnita Watts |
| University Hospitals Bristol and Weston NHS Foundation | St Michaels Hospital | Dr Rachel Liebling | chel Liebling Amy Hanningto |
| University Hospitals Coventry and Warwickshire NHS Trust | | Dr Prakash Satodia | Frankie Brewer |
| University Hospitals of Derby and Burton NHS Foundatio | Queen’s Hospital, Burton | Jane Radford | Claire Prince |
| University Hospitals of Derby and Burton NHS Foundatio | Royal Derby Hospital | Mark Chester | Sarah Miller |
| University Hospitals of Leicester NHS Trust | Leicester Royal Infirmary | Manjiri Khare | Molly Patterson |
| University Hospitals of Leicester NHS Trust | Leicester General Hospital | Dr Manjiri Khare | |
| University Hospitals of North Midlands NHS Trust | Royal Stoke Hospital | Dr Pendee Wu | Anna O'Rourke |
| University Hospitals Plymouth NHS Trust | Derriford Hospital | Sherry Halawa | Heidi Hollands |
| Walsall Healthcare NHS Trust | | Donna Perkins | Vikki Cope |
| Warrington and Halton Teaching Hospitals NHS Foundat | Warrington Hospital | Dr Rita Arya | Lindsay Roughley |
| West Hertfordshire Hospitals NHS Trust | Watford General Hospital | Dr Sankara Narayanan | Dr Uliana Durnea |
| West Suffolk NHS Foundation Trust | West Suffolk Hospital | Barkha Sinha | Lucy Maudlin |
| Western Sussex Hospitals NHS Foundation Trust | St Richard’s Hospital | Emma Meadows | Laura Riddles |
| Western Sussex Hospitals NHS Foundation Trust | Worthing Hospital | | Viv Cannons |
| Wirral University Teaching Hospital NHS Foundation Tru | Arrowe Park Hospital | Julie Grindey | |
| Worcestershire Acute Hospitals NHS Trust | Worcester Royal Hospital | Jessie Brain | Kate Townsend |
| Wrightington, Wigan and Leigh NHS Foundation Trust | Wrightington Hospital | Dr Amit Verma | Claire Williams |
| Wrightington, Wigan and Leigh NHS Foundation Trust | Royal Albert Edward Infirmary | | |
| Wye Valley NHS Trust | Hereford County Hospital | Emma Collins | Melanie Evans |
| Yeovil District Hospital NHS Foundation Trust | | Mr Ahmar Shah | DIANE WOOD |
| York Teaching Hospital NHS Foundation Trust | Scarborough Hospital | Bhavna Pandey | Kerry Elliott |
| York Teaching Hospital NHS Foundation Trust | York Hospital | Robin Hughes | Joanne Ingham |

n
