## Supplementary material for "Pregnancy and neonatal outcomes of COVID-19 – co-reporting of common outcomes from the PAN-COVID and AAP SONPM registry": PAN COVID global investigators

| Hospital | Site | Country | PI or Site lead | Coordinator |
| --- | --- | --- | --- | --- |
| Tongji Medical College of HUST | Union Hospital | China | Mingxing XIE | Li ZHANG |
| Hellenic Society for Maternal Fetal Medicine |  | Greece | Angeliki Gerede MD, | Prof. Apostolos Mamopoulos MD, PhD |
| University Hospital of Ioannina | Neonatal Unit | Greece | Dimitrios Rallis | Maria Fintzou |
| University of Genoa |  | Italy | Fabio Barra | Simone Ferrero |
| Spedali Civili di Brescia | Spedali Civili di Brescia | Italy | Federico Prefumo | Roberta Castellani |
| Government Medical College Kathua Jammu and Kashmir | Associated hospitals and peripheral | India | Dr Pallavi Sharma | Dr Anil Mehta |
| DIVAKARS SPECIALITY HOSPITAL |  | India | Dr Hema Divakar | Dr Poorni Narayanan |
| Universitas Airlangga Hospital | Dr. Soetomo Academic Medical Center | Indonesia | Ernawati Darmawan |  |
| Universitas Airlangga Hospital | Universitas Airlangga Hospital | Indonesia | M.I Aldika Akbar | Khanisyah Erza Gumilar |

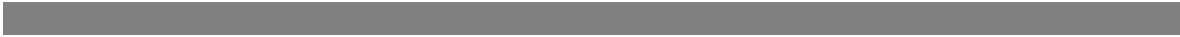
