## Supplementary material for "Pregnancy and neonatal outcomes of COVID-19 – co-reporting of common outcomes from the PAN-COVID and AAP SONPM registry": AAP SONPM investigators

**AAP SONPM National Registry for Surveillance and Epidemiology of Perinatal COVID-19 Infections Participating Centers and Staff:**

From the Department of Pediatrics, Albany Medical Center, Albany, NY: Debra Tristram, Philip Cook, MD; Department of Pediatrics, Ann & Robert Lurie Children’s Hospital of Chicago, Chicago, IL: Kerri Machut, Molly Schau; Department of Pediatrics, Atrium Health Levine Children’s Hospital, Charlotte, NC: Matthew Saxonhouse, Gail Harris; Department of Pediatrics, Avera McKennan Hospital and University Health Center, Sioux Falls, SD: Maria Barber, Christa Friedrich, Jessica Sundleaf; Department of Pediatrics, Banner University Medical Center Phoenix, Phoenix, AZ: Suma Rao, Gretchen Kopec, Christine Wade; Department of Pediatrics, Baptist Medical Center, San Antonio, TX: Kaashif Ahmad, Melanie Drummond; Department of Pediatrics, Barnes Jewish Hospital, St. Louis, MO: Samuel Julian; Department of Pediatrics, Baystate Health, Springfield, MA: Ruben Vaidya, Rachana Singh; Department of Pediatrics, Boston Children’s Hospital, Boston, MA: Amy O’Connell; Department of Pediatrics, Boston Medical Center, Boston, MA: Margaret Parker, Vishakha Sabharwal, Lisa Tucker, Ruby Bartolome ; Department of Pediatrics, Bridgeport Hospital, Bridgeport, CT: Catherine Buck, Christine Henry, Taryn Zamary; Department of Pediatrics, Brigham and Women’s Hospital, Boston, MA: Mandy Belfort, Kaitlin Drouin, Tina Steele; Department of Pediatrics, Brooke Army Medical Center, Fort Sam Houston, TX: Rasheda Vereen, Caitlin Drumm; Department of Pediatrics, St. Joseph / Candler Hospital, Hinesville, GA: Joseph McLean, Meredith Scaccia; Department of Pediatrics, Children’s Memorial Hermann Hospital, Houston, TX: Michael Chang, Gabriela Del Bianco; Department of Pediatrics, Children’s Hospital of New Orleans, New Orleans, LA: Jessica Patrick-Esteves, Peter Joslyn, Christy Mumphrey; Department of Pediatrics, Children’s Hospital of Philadelphia, Philadelphia, NY: David Munson, Dustin Flannery, Madeline Pfeifer; Department of Pediatrics, Children’s Hospital of Richmond at Virginia Commonwealth University, Richmond, VA, Karen Hendricks-Munoz, Russell Moores, Karen Fabian; Department of Pediatrics, Children’s Hospital University of Oklahoma Heath Science Center, Oklahoma City, OK: Patricia Williams, Mike McCoy, Shannon Wilson; Department of Pediatrics, Wilmington Hospital – ChristianaCare, Wilmington, DE: Ursula Guillen, Amy Mackley; Department of Pediatrics, CHRISTUS Santa Rosa Hospital – Westover Hills, San Antonio, TX: Kaashif Ahmad, Melanie Drummond; Department of Pediatrics, Columbia University Irving Medical Center, New York, NY: Donna Garey, Caitlin Ehret; Department of Pediatrics, Connecticut Children’s Medical Center, Hartford, CT: Annmarie Golioto, Nancy Cyr; Department of Pediatrics, Denver Health & Hospital Authority, Denver, CO: Patricia Hagen, Rachel Pena; Department of Pediatrics, Duke University Health System, Durham, NC: Kristin Weimer, Melissa Babilonia-Rosa, Mandy Marion; Department of Pediatrics, East Jefferson General Hospital, Metarie, LA: Jessica Patrick-Esteves, Peter Joslyn, Christy Mumphrey; Department of Pediatrics, Einstein Medical Center Philadelphia, Philadelphia, PA: Agnes Salvador, Gail Cameron; Department of Pediatrics, Emory Decatur Hospital, Decatur, GA: Michelle Pratt, Yvonne Loggins; Department of Pediatrics, Emory Johns Creek Hospital, Johns Creek, GA: Lisa Martin, Yvonne Loggins; Department of Pediatrics, Emory University Hospital Midtown, Atlanta, GA: Ravi Patel, Yvonne Loggins; Department of Pediatrics, Sidney & Lois Eskenazi Hospital, Indianapolis, IN: Gregory Sokol, Leah Engelstad, Hannah Rakow; Department of Pediatrics, Holy Spirit Medical Center, Camp Hill, PA: James A. Cook, Karena Moran; Department of Pediatrics, Geisinger Medical Center, Danville, PA: James A. Cook, Karena Moran; Department of Pediatrics, Geisinger Wyoming Valley Medical Center, Wilkes-Barre, PA: James A. Cook, Karena Moran; Department of Pediatrics, George Washington University Hospital, Washington, D.C.: Ashley Sherwood; Department of Pediatrics, Golisano Children’s Hospital of Southwest FL, Fort Meyers, FL: William Liu; Department of Pediatrics, Good Samaritan Hospital, San Jose, CA: Pui Y. Lai, Rupal Patel, Loc Le; Department of Pediatrics, Good Samaritan Hospital and Medical Center, West Islip, NY: Donna Celetano, Carol Flaim; Department of Pediatrics, Good Samaritan Medical Center, Lafayette, CO: Michelle Feinberg, Laura Celvenger, Kelly Allen; Department of Pediatrics, Grady Medical Center, Atlanta, GA: Ravi Patel, Yvonne Loggins, Colleen Mackie; Department of Pediatrics, Greenwich Hospital, Greenwich, CT: Catherine Buck, Christine Henry, Taryn Zamary; Department of Pediatrics, Hackensack University Medical Center, Hackensack, NJ: Nicole Spillane, Krsytyna Toczylowski; Department of Pediatrics, Henry Ford Hospital, Detroit, MI: Sudhakar Ezhuthachan, Heather Abraam; Department of Pediatrics, Hoag Memorial Hospital Presbyterian, New Port Beach, CA: Sophia Jones, Gazelle Bahramianfard; Department of Pediatrics, Holy Cross Hospital, Silver Springs, MD: Isabelle Von Kohorn, Karla Rondon; Department of Pediatrics, Hospital of Central Connecticut, New Britain, CT: Juliann Sheehan; Department of Pediatrics, Hospital of the University of Pennsylvania, Philadelphia, PA: Elizabeth Foglia, Dustin Flannery, Madeline Pfeifer; Department of Pediatrics, Hackensack University Medical Center Mountainside, Hackensack, NJ: Jonathan Mintzer, Antoine Alexandra Lespinasse; Department of Pediatrics, Hutzel Women’s Hospital, Detroit, MI: Sanket Jani, Monica Bajaj, Jorge Lua, Shanita Binyard; Department of Pediatrics, IU Health Methodist Hospital, Indianapolis, IN: Gregory Sokol, Leah Engelstad, Hannah Rakow; Department of Pediatrics, Jack D. Weiler Hospital, Bronx, NY: Thomas Havranaek, Magdy El-Hennawy; Department of Pediatrics, Jackson-Madison County General Hospital, Jackson, TN: Josefina Go, Laura Richards; Department of Pediatrics, James & Connie Maynard Children’s Hospital, Greenville, NC: Kelly Bear, Sherri Moseley; Department of Pediatrics, John R. Oishei Children’s Hospital, Buffalo, NY: Praveen Chandrasekharan, Emily Li, Jennifer Donato; Department of Pediatrics, Johns Hopkins University, Baltimore, MD: Pamela Donohue, Jennifer Shepard; Department of Pediatrics, Department of Pediatrics, Kaiser Permanente San Francisco Medical Center, San Francisco, CA: Michael Kuniewicz, Allen Fischer, Eileen Walsh, Suyi Zhu; Department of Pediatrics, Kaiser Permanente Modesto Medical Center, Modesto, CA: Michael Kuniewicz, Allen Fischer, Eileen Walsh, Suyi Zhu; Department of Pediatrics, Kaiser Permanente Oakland Medical Center, Oakland, CA: Michael Kuniewicz, Allen Fischer, Eileen Walsh, Suyi Zhu; Department of Pediatrics, Kaiser Permanente Roseville Medical Center, Roseville, CA: Michael Kuniewicz, Allen Fischer, Eileen Walsh, Suyi Zhu; Department of Pediatrics, Kaiser Permanente San Jose Medical Center: Michael Kuniewicz, Allen Fischer, Eileen Walsh, Suyi Zhu; San Jose, CA; Department of Pediatrics, Kaiser Permanente San Leandro Medical Center, San Leandro, CA: Michael Kuniewicz, Allen Fischer, Eileen Walsh, Suyi Zhu; Department of Pediatrics, Kaiser Permanente Santa Clara Medical Center, Santa Clara, CA: Michael Kuniewicz, Allen Fischer, Eileen Walsh, Suyi Zhu; Department of Pediatrics, Kaiser Permanente Santa Rosa Medical Center, Santa Rosa, CA: Michael Kuniewicz, Allen Fischer, Eileen Walsh, Suyi Zhu; Department of Pediatrics, Kaiser Permanente South Sacramento Medical Center, Sacramento, CA: Michael Kuniewicz, Allen Fischer, Eileen Walsh, Suyi Zhu; Department of Pediatrics, Kaiser Permanente Vacaville Medical Center, Vacaville, CA: Michael Kuniewicz, Allen Fischer, Eileen Walsh, Suyi Zhu; Department of Pediatrics, Kaiser Permanente Vallejo Medical Center, Vallejo, CA: Michael Kuniewicz, Allen Fischer, Eileen Walsh, Suyi Zhu; Department of Pediatrics, Kaiser Permanente Walnut Creek Medical Center, Walnut Creek Medical Center, CA: Michael Kuniewicz, Allen Fischer, Eileen Walsh, RN, MPH, Suyi Zhu; Department of Pediatrics, Department of Pediatrics, Kapi’olani Medical Center for Women & Children, Honolulu, HI: Sheree Kuo, Micah Tong; Department of Pediatrics, UK Kentucky Children’s Hospital, Lexington, KY: John Bauer, Beth McKinney-Whitlock, Susan DeGraff; Department of Pediatrics, Lawrence & Memorial Hospital, New London, CT: Catherine Buck, Christine Henry, Taryn Zamary; Department of Pediatrics, M Health Fairview Lakes Medical Center, Wyoming, MN: Cheryl Gale, Jenna Wassenaar; Department of Pediatrics, M Health Fairview Ridges Hospital, Burnsville, MN: Chery Gale, Jenna Wassenaar; Department of Pediatrics, M Health Fairview Southdale Hospital, Edina, MN: Cheryl Gale, Jenna Wassenaar; Department of Pediatrics, Maine Medical Center, Portland, ME: Alan Picarillo; Department of Pediatrics, Maria Fareri Children’s Hospital, Valhalla, NY: Edmund La Gamma, Shetal Shah, Clare Giblin; Department of Pediatrics, Massachusetts General Hospital, Boston, MA: Jessica Shui; Department of Pediatrics, Mayo Clinic, Rochester, MN: Ellen Bendel-Stenzel, Kelly Haines; Department of Pediatrics, Mease Countryside Hospital, Safety Harbor, FL: Jenelle Ferry, Whitney Eldridge; Department of Pediatrics, MedStar Franklin Square Medical Center, Rossville, MD: Siva Subramanian, Tiffany Spriggs; Department of Pediatrics, MedStar Harbor Hospital, Baltimore, MD: Siva Subramanian, Tiffany Spriggs; Department of Pediatrics, MedStar Montgomery Medical Center,
Olny, MD: Siva Subramanian, Tiffany Spriggs; Department of Pediatrics, MedStar Southern Maryland Hospital Center, Clinton, MD: Siva Subramanian, Tiffany Spriggs; Department of Pediatrics, MedStar Washington Hospital Center, Washington, D.C.: Siva Subramanian, Tiffany Spriggs; Department of Pediatrics, UnityPoint Health – Meriter Hospital, Madison, WI: Elizabeth Goetz, Jamie Limjoco, Nina Menda; Department of Pediatrics, Methodist Hospital, San Antonio, TX: Kaashif Ahmad, Katy Kohlleppel; Department of Pediatrics, Methodist Metropolitan, San Antonio, TX: Kaashif Ahmad, Katy Kohlleppel; Department of Pediatrics; Methodist Hospital Stone Oak, San Antonio, TX: Kaashif Ahmad, Katy Kohlleppel; Department of Pediatrics, The MetroHealth System in Cleveland, Cleveland, OH: Prabhu Parimi, Cassandra Smith; Department of Pediatrics, University of Michigan Hospital, Ann Arbor, MI: Rebecca Vartanian, Diane White, Rachael Pace; Department of Pediatrics, Middlesex Health, Middletown, CT: Cliff O’Callahan, Laura Pittari; Department of Pediatrics, Monument Health, Rapid City, SD: Kim Balay, Nan Fitzgerald, Tara O’Leary; Department of Pediatrics, Morristown Medical Center, Morristown, NJ: Caryn Peters; Department of Pediatrics, Morton Plant Hospital, Clearwater, FL: Jenelle Ferry, Whitney Eldridge; Department of Pediatrics, Newton Medical Center, Newton, NJ: Caryn Peters; Department of Pediatrics, Newton-Wellesley Hospital, Newton, MA: Silvia Patrizi; Department of Pediatrics, North Oaks Medical Center, Hammond, LA: Jessica Patrick-Esteve, Peter Joslyn, Christy Mumphrey; Department of Pediatrics, Northeast Baptist Hospital, San Antonio, TX: Kaashif Ahmad, Melanie Drummond; Department of Pediatrics, Northeast Georgia Medical Center, Gainesville, GA: Bridgette Schulman, Aubrey Williams; Department of Pediatrics, Northport Medical Center, Northport, AL: Guillermo Godoy, Carla Kapera; Department of Pediatrics, NorthShore University Health System, Evanston, IL: William MacKendrick, Sue Wolf; Department of Pediatrics, Northside Hospital Atlanta, Atlanta, GA: Mike Hinkes, Katrina Grier, Janna Benston; Department of Pediatrics, Northwestern Medicine Center DuPage Hospital, Winfield, IL: Rita Brennan; Department of Pediatrics, Tisch Hospital, New York, NY: Michelle Vaz, Sourabh Verma, Freda Auyeung; Department of Pediatrics, Oregon Health & Science University, Portland, OR: Dmitry Dukhovny, Monica Rincon, Milica Ivanovic; Department of Pediatrics, OSF Little Company of Mary Medical Center, Evergreen Park, IL: Gretchen Kopec, Michele Astle; Department of Pediatrics, OSF Sacred Heart Medical Center, Danville, IL: Gretchen Kopec, Michele Astle; Department of Pediatrics, OSF St. Anthony’s Health Center, Alton, IL: Gretchen Kopec, Michele Astle, MSN; Department of Pediatrics, St. Mary Medical Center, Galesburg, IL: Gretchen Kopec, Michele Astle, MSN; Department of Pediatrics, Overland Medical Center, Summit, NJ: Kathleen Weatherstone; Department of Pediatrics, Overlook Medical Center, Summit, NJ: Caryn Peters; Department of Pediatrics, Pennsylvania Hospital, Philadelphia, PA: Karen Puopolo, Dustin Flannery, Madeline Pfiefer; Department of Pediatrics, Peyton Mannimakerng Children’s Hospital, Indianapolis, IN: Markus Tauscher, Zenaida Tauscher; Department of Pediatrics, PIH Health Good Samaritan Hospital, Los Angeles, CA: Manoj Biniwale, Rangasamy Ramanathan; Department of Pediatrics, PIH Health Whittier Hospital, Wittier, CA: Liesbeth Maggiotto, Fred Shum, Lisa Batistelli; Department of Pediatrics, Platte Valley Medical Center, Brighton, CO: Sadie Houin, Erin Jones, Kelly Allen; Department of Pediatrics, Pomona Valley Hospital Medical Center, Pomona, CA: Wang-Dar Sun, Hellen Rodriguez, Kenna Schnaar; Department of Pediatrics, UCHealth Poudre Valley Hospital, Fort Collins, CO: Clyde Wright, Jessica Scott, Wendy Barrett; Department of Pediatrics, ProMedica Toledo Hospital, Toledo, OH: Howard Stein, Jessica Shoemaker; Department of Pediatrics, Providence St. Vincent Medical Center, West-Haven Sylvan, OR: Joe Kaempf, Chiayi Chen, Nicole Tipping; Department of Pediatrics, Rady Children’s Hospital, San Diego, CA: Laurel Moyer, Sarah Lazar, Jordan Bui; Department of Pediatrics, Randall Children’s Hospital at Legacy Emmanuel, Portland, OR: Howard Cohen, Kristin Hickey, Lori Keeth; Department of Pediatrics, Regional One Women & Newborn Services, Memphis, TN: Vineet Lamba, Gail Camp; Department of Pediatrics, Riley Hospital for Children, Indianapolis, IN: Gregory Sokol, Leah Engelstad, Hannah Rakow; Department of Pediatrics, Rush Copely Medical Center, Aurora, IL: Melissa Knapik; Department of Pediatrics, Rush University Medical Center, Chicago, IL: Andrew Berenz, Megan Gross; Department of Pediatrics, Saint Barnabus Medical Center, Chicago, IL: Kwanchai Chan, Deborah Ann Cialfi; Department of Pediatrics, Saint Joseph Hospital, Denver, CO: Alfonso Pantoja, Corrie Alonzo, Allie Wildenstein, Kelly Allen; Department of Pediatrics, Salem Health, Salem, Oregon: Christopher Traudt; Department of Pediatrics, Santa Clara Valley Medical Center, Fruitdale, CA: Priya Jegatheesan, Angela Huang; Department of Pediatrics, Silver Cross Hospital, Lennox, IL: Colleen Malloy Stover, Marilyn Paolella; Department of Pediatrics, Slidell Memorial Hospital, Slidell, LA: Jessica Patrick-Esteve, Peter Joslyn, Christy Mumphrey; Department of Pediatrics, Sparrow Hospital, Lansing, MI: Said Omar, Cheryl Abernathy, Sara Hackett; Department of Pediatrics, Spartanburg Medical Center, Spartanburg, SC: Imelda Uy; Department of Pediatrics, SSM Health DePaul Hospital, St. Louis, IL: Justin Josephsen, Melissa Hawkins; Department of Pediatrics, SSM Health St. Mary’s Health – Audrain, Mexico, IL: Justin Josephsen, Melissa Hawkins; Department of Pediatrics, St. Joseph’s Hospital, Tampa, FL: Jenelle Ferry, Whitney Eldridge; Department of Pediatrics, St. Vincent Healthcare, Billings, MT: Alison Rentz; Department of Pediatrics, St. Vincent’s Mercy Medical Center, Toledo, OH: Gagandeep Brar, Kelly Parker, Christine Calcamuggio; Department of Pediatrics, St. Bernards Medical Center, Jonesboro, AR: Enrique Gomez, Hailey McNew, Christal Steen; Department of Pediatrics, HSHS St. John’s Children’s Hospital, Springfield, IL: Beau Batton, Allison Spenner; Department of Pediatrics, St. Joseph’s Medical Center, Patterson, NJ: Linda Skroce; Department of Pediatrics, St. Joseph’s Women’s Hospital, Tampa, FL: Jenelle Ferry, Whitney Eldridge; Department of Pediatrics, St. Joseph Hospital, Orange, CA: Renuka Kar, Maria Ransil; Department of Pediatrics, St. Jude Medical Center, Fullerton, CA: Lavonne Sheng, Terry Zeilinger; Department of Pediatrics, St. Louis Children’s Hospital, St. Louis, MO: Samuel Julian, Laura Linneman; Department of Pediatrics, St. Luke’s Health System, Boise, ID: Jennifer Merchant, Nichele Parks; Department of Pediatrics, Tampa General Hospital – USF, Tampa, FL: Tara M. Randis, Marcia Kneusel; Department of Pediatrics, Temple University Hospital, Philadelphia, PA: Heidi Taylor, Sruthi Polavarapu; Department of Pediatrics, The Children’s Hospital of San Antonio, San Antonio, TX: Kaashif Ahmad, Melanie Drummond; Department of Pediatrics, Valley Hospital Medical Center, King of Prussia, PA: Christiana Farkouh-Karoleski, Peggy Bischoff; Department of Pediatrics, Touro Infirmary LCMC Health, New Orleans, LA: Jessica Patrick-Esteve, Peter Joslyn, Christy Mumphrey; Department of Pediatrics, Mercy Health Saint Mary’s, Grand Rapids, MI: Steven Gelfand, Stacy Smith; Department of Pediatrics, Tufts Medical Center, Boston, MA: Jill L. Maron, Taysir Mahmoud; Department of Pediatrics, Tulane Lakeside Hospital, Metarie, LA: Elizabeth Lindsay, Leslie Smith, Monique Diles; Department of Pediatrics, UC Health Medical Center of the Rockies Trauma Center, Loveland, CO: Clyde Wright, Jessica Scott, Wendy Barrett; Department of Pediatrics, UC Irvine Medical Center, Orange, CA: Cherry Uy, Pam Aron-Johnson; Department of Pediatrics, UC Health Memorial Center, Colorado Springs, CO: Clyde Wright, Jessica Scott, Sue Townsend; Department of Pediatrics, UC Health Memorial Hospital North, Colorado Springs, CO: Clyde Wright, Jessica Scott, Sue Townsend; Department of Pediatrics, UC San Diego Health, San Diego, CA: Ericka Fernandez, Sarah Lazar, Melanie Crabtree; Department of Pediatrics, UF Health Jacksonville, Jacksonville, FL: Josef Cortez, Ashley Maddox; Department of Pediatrics, UF Health Shands Gainesville, Gainesville, FL: David Burchfield, Livia Sura; Department of Pediatrics, U Mass Memorial Health Medical Center, Worcester, MA: Katherine Sullivan, Heather White, Archana Kalyanasundaram; Department of Pediatrics, University of North Carolina Children’s Hospital (NICU/PICU), Chapel Hill, NC: Genevieve Taylor, Jennifer Talbert; Department of Pediatrics, United Health Services Hospitals, Inc., Binghamton, NY: Paula Farrell, Marybeth Culp, Terri Peters; Department of Pediatrics, University Hospital in San Antonio, San Antonio, TX: Joseph B. Cantey, Diana Guerra; Department of Pediatrics, University Hospitals Cleveland Medical Center, Cleveland, OH: Michele C. Walsh, Anna Maria Hibbs, Nancy Newman, Riddhi Desai; Department of Pediatrics, University of Arkansas for Medical Science Medical Center, Little Rock, AR: Richard Whit Hall, Dalton Janssen; Department of Pediatrics, University of Colorado Hospital, Aurora, CO: Clyde Wright, Jessica Scott, Kathleen Hannan; Department of Pediatrics, University of Illinois Hospital & Health Science System, Chicago, IL: De-Ann Pillers, Zaynab Kadhem; Department of Pediatrics, University of Iowa Stead Family Medical Center, Iowa City, IA: Timothy Elgin, Karen Johnson; Department of Pediatrics, M Health Fairview University of Minnesota Masonic Children’s Hospital, Minneapolis, MN: Cheryl Gale, Jenna Wassenar; Department of Pediatrics, University of Mississippi Medical Center, Jackson, MS: Jagdish Desai, Heather Williams, Aurora Diaz; Department of Pediatrics, University of Pittsburg Medical Center, Pittsburg, PA: Toby Yanowitz, Victoria D’Orto; Department of Pediatrics, UR Medicine – Strong Memorial Hospital, Rochester, NY: Kristin Scheible, Mallory Prideaux, Rachel Jones, Tanya Scalise; Department of Pediatrics, University of Virginia Children’s Hospital, Charlottesville, VA: Jonathan Swanson, Monika Thielen; Department of Pediatrics, Valley Medical Center – UW Medical Center, Renton, WA: Christina Long, Toby Cohen; Department of Pediatrics, University of Tennessee Medical Center Knoxville, Knoxville, TN: Courtney Gutman: Department of Pediatrics, Vanderbilt University Medical Center, Nashville, TN: Hendrik Weitkamp, Theresa Rogers; Department of Pediatrics, Virtua Memorial Hospital, Mount Holly, New Jersey: Sarvin Ghavam, Christine Catts, Jonathan Snyder; Department of Pediatrics, Virtua Voorhees Hospital, Voorhees Township, NJ: Sarvin Ghavam, Christine Catts, Jonathan Snyder; Department of Pediatrics, Brenner Children’s Hospital – Wake Forest Baptist Health, Winston-Salem, NC: Jeffrey S Shenberger, Cobi Ingram; Department of Pediatrics, WakeMed Health Raleigh Campus, Raleigh, NC: Stephen Kicklighter, Donna White; Department of Pediatrics, Wellspan York Hospital, York, PA: Elias Abebe, Michael Goodstein, Michelle Eppinger; Department of Pediatrics, Winchester Medical Center, Winchester, VA: Edward I. Lee; Department of Pediatrics, Wolfson Children’s Hospital, Jacksonville, FL: Colby Day-Richardson, Ashley Maddox; Department of Pediatrics, Woman’s Hospital, Baton Rouge, LA: Steven B. Spedale; Department of Pediatrics, Yale New Haven Hospital, New Haven, CT: Catherine Buck, Christine Henry, Taryn Zamary.
