## Supplementary material for "Pregnancy and neonatal outcomes of COVID-19 – co-reporting of common outcomes from the PAN-COVID and AAP SONPM registry": PAN COVID data fields

**Welcome to the PAN-COVID register** PAN-COVID

| | | | |
|---|---|---|---|
| | **Please circle your answers** | | |
| **CONSENT** | Has the participant provided verbal informed consent? | Yes | No |
| | Date provided consent | | |
| **DOB** | What is the participant's date of birth? | | |
| **NHS/ID no.** | What is the participant's hospital number (e.g. NHS or CHI)? | | |
| **EDD** | Does the participant have an **expected** date of delivery **based on ultrasound**? | Yes | No |
| | If yes, please provide the **expected** delivery date based on ultrasound | | |
| | If no, please provide the date of participant's last menstrual period | | |
| **BMI** | What is the participant's BMI? | | Date measured/calculated: |
| **SMOKE** | Does the participant currently smoke cigarettes or tobacco? | Yes, participant smokes | |
| | | No, participant has never smoked | |
| | | No, participant used to smoke but stopped before this pregnancy | |
| | | No, participant stopped after they knew they were pregnant | |
| **ETHNICITY** | What is the participant's ethnicity? | European / North American | |
| | | Middle East | |
| | | Northern Africa | |
| | | Africa south of Sahara / Caribbean | |
| | | Indian subcontinent | |
| | | SE Asia | |
| | | South - Middle America | |
| | | Other | |
| **COVID/ SARS-CoV-2 infection** | Please describe the status of participant's COVID-19 diagnosis? | | Date of test or first symptom onset |
| | | Suspected | |
| | | Confirmed | |
| | | Negative test | |
| | If suspected, please circle all symptoms which apply | Fever | |
| | | New, persistent cough | |
| | | Anosmia | |
| | | Myalgia | |
| | | Diarrhoea | |
| | | Shortness of breath | |
| | | Fatigue | |
| | | Loss of appetite | |
| | | None of the above | |
| **Medications** | | | |
| | Aspirin | Yes | No |
| | Progesterone | Yes | No |
| | Immunosuppresion | Yes | No |
| | Low molecular weight heparin | Yes | No |
| | Any pregnancy vitamins | Yes | No |
| | Other | Yes | No |
| | None | Yes | No |
| **Medical History** | | | |
| | Chronic hypertension | Yes | No |
| | Pregnancy-induced hypertension | Yes | No |
| | Respiratory disease | Yes | No |
| | Cardiovascular disease | Yes | No |
| | Renal disease | Yes | No |
| | Autoimmune disease | Yes | No |
| | Pre-existing diabetes | Yes | No |
| | Gestational diabetes | Yes | No |
| | Other | Yes | No |
| | None | Yes | No |
| **Gravidity** | Please list outcome of previous pregnancies (miscarriage/pregnancy loss/livebirth, gestation, birthweight, neonatal death) | | |
| 1 | | | |
| 2 | | | |
| 3 | | | |
| 4 | | | |
| 5 | | | |
| 6 | | | |
| 7 | | | |
| 8 | | | |
| **Pregnancy details** | | | |
| | Please provide the number of fetuses the participant is carrying? | | |
| | Does the participant have pre-eclampsia? | Yes | No |
| | Does the participant have eclampsia? | Yes | No |
| | Does the participant have fetal growth restriction? | Yes | No |
| | If has fetal growth restriction, please circle all that apply | Abdominal circumference or estimated fetal weight < 3rd centile | |
| | | Umbilical artery or uterine artery PI > 95th centile | |
| | | Abdominal circumference or estimated fetal weight reduced from 20/40 scan and crossed 40 centiles | |
| | | Cerebro-umbilical ratio < 5th centile | |
| | Fetal structural malformation(s) present on ultrasound? | Yes | No |
| | If yes, please circle all that apply | | Head |
| | | | Brain |
| | | | Central Nervous System |
| | | | Heart |
| | | | Limb |
| | | | Gastroenteritis |
| | | | Urinary |
| | | | Genital |
| | Date of the participant's delivery? | | |
| | Was the participant's labour induced? | | |

| | | | |
|---|---|---|---|
| What was the indication for delivery? | | | |
| What was the outcome of delivery? | | Miscarriage | |
| | | Termination of pregnancy | |
| | | Livebirth | |
| | | Intra-uterine death/stillbirth (>22+6 weeks gestation) | |
| If yes to miscarriage, did the woman have a previous ultrasound scan? | Yes | No | |
| If yes, please circle the diagnosis at the previous scan | | Viable intra-uterine pregnancy | |
| | | Pregnancy unknown viability | |
| | | Pregnancy unknown location | |
| Birthweight baby 1 (g) | | | |
| Birthweight baby 2 (g) | | | |
| Did COVID-19 lead to the participant requiring any of the following? Please circle all that apply and give dates | | | |
| | ***Date started*** | ***Date stopped*** | |
| Non invasive ventilation | | | |
| Intubation and ventilation | | | |
| Did the participant die? | Yes | No | If yes, date: |
| If Yes, please circle the presumed cause of death | | COVID-19 | |
| | | Pregnancy related | |
| | | Other | |
| If the participant had a livebirth, please provide the NHS number (or ID number) for the baby/babies delivered | Baby 1 | | |
| | Baby 2 | | |
| Please provide the gender of the baby/babies | Baby 1 | Male | Female |
| | Baby 2 | Male | Female |
| Congenital malformation(s) present at delivery? | Yes | No | |
| If yes, please circle all that apply (if >1 baby, please indicate which baby affected) | Head | | |
| | Brain | | |
| | Central Nervous System | | |
| | Heart | | |
| | Limb | | |
| | Gastroenteritis | | |
| | Urinary | | |
| | Genital | | |
| What was the APGAR score at 5 minutes? | | | |
| If >1 baby, what was the APGAR score of the second neonate at 5 minutes? | | | |
| Baby(s) separated from the participant immediately after delivery? | Yes | No | |
| If yes, how many days were they separated? | | | |
| Has the baby(s) received breastmilk from the participant? | | | |
| Has the baby(s) been tested for SARS-COV-2? | Yes | No | |
| If Yes, please indicate sample(s), date(s) and result(s) | ***Date*** | ***Sample*** | ***Result*** |
| Did the participant's neonate(s) experience any complications? | Yes | No | |
| If yes, please circle any that apply | Transient tachypnea of newborn | | |
| If >1 baby, please indicate which baby/babies affected | Respiratory distress syndrome | | |
| | Pneumonia | | |
| | None of the above | | |
| Did the participant's baby/babies die? | Yes | No | |
| If yes, what was the date of death | Baby 1 | | |
| | Baby 2 | | |
| If yes, what was the suspected cause of death | Baby 1 | | |
| | Baby 2 | | |
| Was the participant's baby/babies re-admitted to hospital at any point between date of delivery and 28 days old? | Yes | No | |
| If yes, what was the date of re-admission? | | | |
| If yes, what was the date the baby was discharged? | | | |
