## Supplementary material for "Pregnancy and neonatal outcomes of COVID-19 – co-reporting of common outcomes from the PAN-COVID and AAP SONPM registry": AAP SONPM data dictionary

### NATIONAL REGISTRY FOR SURVEILLANCE AND EPIDEMIOLOGY OF PERINATAL COVID-19 INFECTION

Case definitions of the mother/infant dyads

For inborn infants (born at your reporting institution):

A pregnant woman (1) who is known to have had virologic testing positive for SARS-CoV-2 within 14 days prior to delivery or (2) who is admitted as a PUI and whose peri-delivery testing obtained through 72 hours after delivery Is reported to be positive for SARS-CoV-2; and her infant(s).

For outborn infants (born at a referring hospital and transferred to your institution):

A mother who is known to have had virologic testing positive for SARS-CoV-2 obtained within 14 days prior to through 72 hours after delivery at an outside hospital; and her infant(s) who is (are) transported to your hospital for a higher level of care.

Definitions of data elements

Age of mother
Completed years of age of mother, based on date of birth

Gravidity
The number of times the mother has been pregnant, including the current pregnancy

Race of mother

**Black or African American**
If the biological mother is a person having origins in any of the black racial groups of Africa

**White**
If the biological mother is a person having origins in any of the original peoples of Europe, the Middle East, or North Africa

**Asian**
If the biological mother is a person having origins in any of the original peoples of the Far East, Southeast Asia, or the Indian subcontinent including, for example, Cambodia, China, India, Japan, Korea, Malaysia, Pakistan, the Philippine Islands, Thailand, and Vietnam

**American Indian or Alaska Native**

If the biological mother is a person having origins in any of the original peoples of North and South America (including Central America), and who maintains tribal affiliation or community attachment

**Native Hawaiian or Other Pacific Islander**

If the biological mother is a person having origins in any of the original peoples of Hawaii, Guam, Samoa, or other Pacific Islands

**Other**

If none of the race categories above applies to the biological mother

Ethnicity of mother

**Hispanic**

If the biological mother is a person of Cuban, Mexican, Puerto Rican, South or Central American, or other Spanish culture or origin, regardless of race

**Non-Hispanic**

If the biological mother's ethnicity is not of Hispanic or Latino origin as defined above.

COVID-19 status at delivery **(Must provide a value)**

**Confirmed COVID-19**

If results of virological testing for COVID-19 performed in the 14 days before delivery are known to be positive at the time of delivery

**PUI (Person Under Investigation)**

If the initial positive results of virological testing for COVID-19 obtained from 14 days before delivery to 3 days after delivery first become known after delivery

Interval (days) between date of admission for delivery and date of neonatal birth

Example:

A mother is admitted to your institution on June 1st and delivers on June 3rd. The interval is 2 days.

Note: **If infant was outborn and these data are unavailable, please leave blank**

Interval (days) between **first** positive maternal test for SARS-CoV-2 and date of birth

Examples:

The date of the first positive laboratory result for SARS-CoV-2 is June 1st and the infant is delivered on June 3rd. The interval is +2 days.

The date of birth was June 3rd and the date of the first positive laboratory result for SARS-CoV-2 was June 5th. The interval is -2 days.

Note: **If infant was outborn and these data are unavailable, please leave blank**

*Interval (days) between date of first maternal symptoms c/w COVID-19 infection and date of delivery (Positive number if symptoms occurred before delivery; negative number if symptoms did not appear until after delivery; zero if symptoms first developed on the day of delivery)

Example 1:

A mother first develops symptoms c/w COVID-19 infection on May 1. She is confirmed to be positive for SARS-CoV-2 by PCR test on a specimen obtained on May 4. She gradually recovers from her illness over two weeks. She is admitted with no active symptoms for delivery at term on May 28. Her repeat PCR test on a specimen obtained on May 28 is positive. She delivers a healthy baby on May 29.

The interval between the date of first maternal symptoms (May 1) and the date of delivery (May 29) is 28 days.

Example 2:

If a mother had no symptoms c/w COVID-19 infection at any time during pregnancy or in the first few days after pregnancy, respond with the text entry, “no symptoms”.

*Number of days between the **most recent** maternal positive test and DOB

Example:

A mother first develops symptoms c/w COVID-19 infection on May 1. She is confirmed to be positive for SARS-CoV-2 by PCR test on a specimen obtained on May 4. She gradually recovers from her illness over two weeks. She is admitted with no active symptoms for delivery at term on May 28. Her repeat PCR test on a specimen obtained on May 28 is positive. She delivers a healthy baby on May 29.

The number of days between the most recent positive test (May 28) and the DOB (May 29) is 1 day.

Duration of maternal hospitalization (days)

Example:

A mother is admitted to your institution on June 1st, and discharged on June 20th. The total days of hospitalization is 20 days.

Note: **If infant was outborn and these data are unavailable, please leave blank**

Final maternal disposition status

**Discharged home**

Mother was discharged home from your institution

**Transferred to other facility**

Mother was transferred from your institution to another hospital, either as a step up for acute care, or as a step down for discharge planning

**Expired (provide reason below)**

The mother died before discharge home. If you select this answer, you will be prompted to complete an additional field detailing the cause of maternal death:

**Related to COVID-19**
**Not related to COVID-19**
**Unknown**

Indication for which mother was tested (choose all that apply)

**URI symptoms**

Cough, nasal congestion, sinus congestion, sore throat

**LRI symptoms**

Respiratory distress, cough, wheezing

**Fever**

Defined as measured temperature $\geq$ 37.8° C

**GI symptoms**

Vomiting, diarrhea, nausea

**Myalgia and fatigue**

Muscle pains and aches, loss of energy

**Anosmia/ageusia**

Defined as loss of smell or taste, may be partial or complete

**Contact with SARS-CoV-2 case**

Direct exposure to a known laboratory positive SARS-CoV-2 individual in the 14 days prior to testing

**Travel**

Any travel outside of the US, travelers returning from high risk countries, cruise ship travel, travel to high risk areas domestically where COVID-19 cases are known to be high (i.e. New York City)

**Unknown**

Mother tested for unknown causes

**Other**

Testing performed for a reason not represented in selections above. Please provide detailed explanation in box provided

Note: **If infant was outborn and these data are unavailable, please leave blank**.

Maternal condition before admission for delivery

**Asymptomatic**

No known signs or symptoms of COVID-19 infection

**Sick at home**

Mother had one or more symptoms of COVID-19 infection, but did not require hospital admission. (If selected, you will be prompted to identify symptoms which are the same as those listed under "Indication for which mother was tested"

**Required hospitalization for SARS-CoV-2 before delivery (anytime within 14 days prior to delivery)**

Mother was admitted to a hospital within 14 days prior to delivery for an illness known to be or later identified as COVID-19 infection (If selected, you will be prompted to identify maternal therapies: choose all that apply).

**IV fluids**

**Supplemental O2**

**CPAP**

**Mechanical ventilation**

**ECMO**

**Other**

Provide description in the textbox.

Days of illness before delivery (number)

Example:

A mother begins to feel ill on June 1st, and the mother delivers on June 20t. The days of illness are 20.

Note: **If infant was outborn and these data are unavailable, please leave blank**

Labor (choose one)

**Spontaneous**

Labor onset occurs without cervical ripening or intravenous agents.

**Augmented**

Stimulation of the uterus to increase the frequency, duration and intensity of contractions after the onset of spontaneous labor.
If you select augmented or induced, additional questions will populate regarding maternal or fetal indications

**Induced**

Stimulation of uterine contractions after spontaneous onset of labor
If you select augmented or induced, additional questions will populate regarding maternal or fetal indications

**None**

Delivery occurred without labor

Route of delivery

**Vaginal**

**C-section**

If you selected spontaneous labor and c-section, additional questions will populate regarding maternal or fetal indications that led to c-section delivery

If labor was augmented or induced OR delivery was by c-section, what was (were) the indication(s)? (choose all that apply)

Maternal indications (choose all that apply)

**Post dates**

Infant not delivered by 41 weeks' gestation

**Preeclampsia**

High blood pressure, hyperreflexia, and proteinuria

**Arrest of descent**

Failure for the head to progress in the second stage after the cervix is completely dilated

**Abruption**

Partial or complete separation of the placenta from the inner wall of the uterus prior to delivery

**Chorioamnionitis**

A clinical diagnosis suggesting inflammation of the amnion and chorion of the placenta, due to bacterial infection

**Concern about worsening maternal condition due to COVID-19 infection should pregnancy continue**

Acute decompensation of mother requiring interventions such as intubation, ventilation, and cardiovascular support

**Other**

Any indication not represented by the causes listed above. Please provide detail in textbox provided.

Fetal indications (choose all that apply)

**Severe intrauterine growth restriction**

Weight estimated to be less than the 10$^{th}$ percentile for the gestational age as per the population growth charts

**Fetal distress**

Concern that the fetus has inadequate oxygen delivery or blood flow. May be evidenced by fluctuations in fetal HR such as late decelerations, variable decelerations, or prolonged bradycardia

**Hydrop fetalis**

Fetal edema, ascites, and/or pleural effusions

**Malpresentation**

When a fetal part other than the head engages maternal pelvis (i.e. a breech presentation)

**Other**

Any indication not represented by the causes listed above. Please provide detail in the textbox provided.

Rupture of Membranes (ROM)

**Spontaneous**

Natural rupture of the amniotic sac

**Artificial**

Rupture accomplished by external procedure.

Duration of ROM (hours)

Length of time in hours from initial rupture of membranes until the delivery of the fetus.

Did mother have negative SARS-CoV-2 testing after the positive test?

**Yes**

If mother had negative SARS-CoV-2 testing after delivery. (If you select yes, then a box will appear asking you to enter the number of days from delivery that mother had the negative test).

Example:

A mother delivered June 1st and on June 3rd had a negative SARS-CoV-2 test. The days would be 2.

**No**

If mother did not have a negative SARS-CoV-2 test after delivery

**Unknown**

If it is unknown whether the mother had a negative SARS-CoV-2 test after delivery

Did mother receive betamethasone before delivery

**Yes**

Mother received at least 1 dose prior to delivery

**No**

Mother did not receive any betamethasone prior to delivery

**Unknown**

It is unknown whether mother received betamethasone prior to delivery

Did mother receive specific treatment for SARS-CoV-2?

**Yes**

Mother received SARS-CoV-2 treatment. (An additional question will populate regarding what drug was given for treatment)

If yes, what medication(s) did mother receive (choose all that apply)

**Chloroquine**

**Hydroxychloroquine**

**Remdesivir**

**Ritonavir**

**Favipiravir**

**Lopinavir**

**Other**
If you select other, enter any other treatments or medications the mother received in the text box.

**No**
Mother did not receive SARS-CoV-2 treatment
.
**Unknown**
It is unknown whether mother received SARS-CoV-2 treatment

**NEWBORN**

Gestational age at birth (to nearest completed weeks)
Please complete to nearest completed gestational week i.e. 36w3d = 36 weeks

Birth weight (g)
Enter the birth weight in grams

Sex
**Male**
**Female**

Apgar at 5 minutes
5 min Apgar score at 5 minutes; if unknown, enter -1

Status at birth
**Liveborn**

**Stillborn**

**Death in delivery room**

Birth multiplicity
**Single**

**Twin A**

**Twin B**

**Higher order (triplet, etc.)**

Resuscitation at birth (check all that apply)
**Drying and stimulation only**
Infant requires on drying and stimulation for resuscitation

**Oxygen**

Infant requires oxygen as part of resuscitation (blow-by only, or with other modalities)

**Positive pressure (CPAP or mask ventilation)**

Infant requires positive pressure to be applied via CPAP or face mask ventilation (may be with or without oxygen).

**Intubation**

Infant requires intubation as part of resuscitation

**Chest compressions**

Infant requires any chest compressions as part of resuscitation

**Epinephrine**

Infant requires the use of epinephrine as part of resuscitation (via endotracheal tube or IV)

**Volume expansion**

Infants requires volume expansion as part of resuscitation (i.e normal saline bolus)

Were mother and infant separated at <u>your</u> hospital at birth (if inborn) or was mother not given visitation rights until she was virologically cleared (if outborn)?

**Yes**

**No**

**Unknown**

Was the infant isolated at <u>your</u> hospital (choose all that apply)?

**Yes – enhanced respiratory droplet**

**Yes – airborne precautions**

**Yes – negative pressure room**

**Yes – other (Provide below)**

If yes, enter other isolation method in textbox.

**No**

**Unknown**

What were the locations at which the infant was cared for at your hospital (choose all that apply)

**Neonatal Intensive care setting**

The infant was cared for in the Neonatal ICU at your institution

**Nursery setting separate from mother**

The infant was cared for separate from mother in the newborn nursery

**Room-in with mother**

The infant and mother were cared for together in the same room

**Negative pressure isolation room**

The infant was cared for in a negative pressure isolation room, (a negatively pressured room with respect to adjacent areas to prevent airborne contaminants / pathogens from drifting to other areas) where Airborne Precautions were in place

**Regular isolation room**

The infant was cared for in a standard isolation room where Respiratory Droplet Precautions were in place

**Other**

The infant was cared for in a setting not previously mentioned above. Please enter other location(s) in textbox.

Duration of neonatal hospitalization at your hospital (days)

Example:

An infant is admitted to your institution on June 1st, and discharged on June 20th. The total days of hospitalization would be 20

Final disposition status

**Discharged home**

The infant is discharged directly home from your institution

**Transferred to another facility**

The infant is transferred from your institution as a step up for acute care, or as a step down for discharge planning

**Expired (provide reason below)**

The infant expired. (If you select expired, you will be prompted to complete an additional field detailing cause of newborn death)

Cause of newborn death –

**Related to COVID-19**

**Not Related to COVID-19**

**Unknown**

Primary reason for neonatal transfer to <u>your</u> hospital

**Clinical illness presumed related to COVID-19 infection**

**Other clinical illness not presumed to be related to COVID-19 infection**

**No clinical illness; transport due to maternal COVID-19 diagnosis**

**N/A (infant was inborn at your hospital)**

Neonatal signs during hospitalization (choose all that apply)

**None**

The infant had no signs or symptoms of concern

**Fever (>37.8° C)**

The infant had at least one measured temperature >37.8° C

**Cough**

**Vomiting/diarrhea**

**Respiratory distress diagnosis** (provide below)

The infant had a diagnosis of respiratory distress (If checked, enter details in textbox)

**Hypotonia**

The infant had decreased muscle tone

Maximal level of respiratory support during hospitalization (select one):

**None**

**Supplemental oxygen**

**CPAP**

**Mechanical ventilation**

**ECMO**

Other diagnoses (choose all that apply)

**None**

**Delayed transition**

The infant had signs of respiratory distress that resolved within the first few hours of life (i.e transient tachypnea)

**Surfactant deficiency**

The infant had a clinical diagnosis of RDS

**Pulmonary hypertension**

The infant had documented pulmonary hypertension on cardiac echo

**Hypotension**

The infant had blood pressures that were treated with volume, pressors, or steroids

**Hypoglycemia**

The infant had documental blood glucose less than 30 mg/dL in the first 24 hours of life and/or less than 45 mg/dL thereafter

**Hypothermia**

The infant has measured core temperature less than 35 C

**Culture-confirmed bacterial sepsis (organism) (Provide below)**

The infant had a positive blood culture that is positive. (If you check this diagnosis, enter the organism(s) in the textbox)

**Microcephaly**

The infant had a head circumference within the first 24 hours of life which is smaller than expected when compared to babies of the same sex and age (2 standard deviations smaller)

**Encephalopathy**

The infant had documented signs of abnormal neurologic function in the first few day of life due to brain injury (i.e. birth asphyxia, cerebral hypoxia, HIE)

**Congenital anomalies (provide below)**

If you select this option, enter anomaly(ies) in the textbox

**Other**

If you select this option, enter information in the textbox

Other administered intensive care (choose all that apply)

**None**

**Antibiotics**

**Antivirals**

If you select this option, enter information in the textbox.

**Intravenous fluids**

.

**Inhaled nitric oxide or prostacyclin**

**Pressors**

If you select this option, choose all that apply

**Dopamine**

**Dobutamine**

**Epinephrine**

**Surfactant**

**Hydrocortisone**

.

**Dexamethasone**

**Therapeutic hypothermia**

The infant received full body or selective head cooling

**ECMO**

**Other**

If you select this option, choose all that apply

**[For Laboratory findings from birth to day 7 (choose all recorded), a table will appear for you to record the value obtained and day of age for your results. For example, if an infant is born on June 1st, the day of birth would be day 0. If that same infant has a lab result on June 3rd, the day of age would be 2.]**

Laboratory findings from birth (day 0) to day 7 (choose all recorded)

**Lowest white count**

**Lowest neutrophil count**

**Lowest lymphocyte count**

**Lowest hemoglobin**

**Highest AST**

**Highest ALT**

**Highest CRP**

Did the newborn have SARS-CoV-2 testing during the first 14 days of life?

Example:
If an infant is born on June $1^{s}$ the day of birth would be day 0. If that same infant has a SARS-CoV-2 test on or prior to June $14^{th}$, then the answer would be yes

**Yes**

**No**

Was the newborn admitted to a higher level of care (e.g. NICU) than a newborn nursery or general hospital ward (for boarding care)?

**Yes**

**No**

Did the mother provide breast milk (choose all that apply) –

**Yes – direct nursing by mother**
The infant latched on and fed at the breast

**Yes – expressed maternal milk fed by mother**
The infant fed milk provided by the mother via breast pump that was bottle fed to the infant

**Yes** – **expressed maternal milk fed by another caregiver**
The infant fed milk provided by the mother via breast pump that was fed to the infant by another caregiver (bottle or feeding tube)

**Yes** – **donor milk**
The infant was fed donor breast milk
.

**No**
The infant did not receive breast milk

**Unknown**

If the infant received mother’s own milk, either by direct breast feeding or by expression, what was the viral testing status of the mother at that time?

**Positive**

The mother’s viral testing for SARS-CoV-2 was documented as positive

**Pending**

The mother’s viral testing is pending and has not been resulted prior to feeding

**Negative**

The mother’s viral testing is documented as negative

**Not tested**

The mother never received viral testing

**Unknown**

The mother’s viral testing status is unknown.

Discharge medications

**None**

**Yes**

If you select yes, enter details in the textbox

Email address of the member of the study team submitting these data

Self-populated during authentication

Assigned sequenced tracking number (e.g., 1, 2, 3, 4 etc. in order of submission)

Please number each of your CRFs in order with cardinal numbers beginning with 1. Only your team will be able to link your site and patient tracking number to the patient MRN, which allows your submission to be de-identified. Please enter mother/infant dyad data according to the chronological order of birth.

When all of your data entry is complete, hit the SUBMIT button.
